# More than the sum of their parts: A Behavioral Tractography analysis of distress wellbeing and context reveals dynamic signatures of psychosis and autism

**DOI:** 10.1101/2025.07.09.25330999

**Authors:** Corrado Sandini, Andrea Imparato, Clémence Feller, Laura Ilen, Natacha Reich, Christopher Graser, Stephan Eliez, Argyris Stringaris, Maude Schneider

## Abstract

Current psychiatric assessments oversimplify mental health phenomena. Ecological-Momentary-Assessment (EMA) captures richer dynamic interactions between psychological and contextual variables, but the resulting complexity currently hinders clinical translation. Here we propose an innovative approach to analyze dynamic clinical data by accounting for variations within and across symptom clusters over time. We term this approach Behavioral-Tractography. We validate Behavioral-Tractography in independent typically-developing samples and in the clinically-challenging differential diagnosis of psychosis genetic vulnerability in 22q11.2 Deletion Syndrome (22q11DS) and idiopathic Autism Spectrum Disorder (ASD). A novel 2-dimensional rationality/valence structure predicted behavioral fluctuations with striking consistency across typically-developing and clinical samples. Characterizing dynamic interactions between variables within such a shared coordinate system, provided an intuitive 3D-view of both transdiagnostic and population-specific behavioral pathways. 22q11DS and ASD presented opposite behavioral dynamics, recapitulating Bayesian models of psychosis and autism, and were further differentiated by pathways mediating emergence and consequences of motivational and social wellbeing.

## Introduction

Mental disorders are a leading cause of disease burden (1). Yet, our understanding of them (2) and our ability to link them to advances in neuroscience (3) remain limited. This limitation may, at least in part, stem from how disorders and symptoms have been assessed (4–6). Traditional psychiatric assessments rely on retrospective reports measuring the average intensity of symptoms—such as delusions, irritability, or avolition—over weeks or months (4). Due to the inherent limitations of subjective recall (7), this approach fails to capture symptom fluctuations occurring over shorter periods, such as hours or days (8), which may carry crucial information for understanding and managing mental health disorders (6, 9). Recent advancements in ambulatory assessment techniques, such as Ecological Momentary Assessment (EMA), have provided a unique window into the dynamics of mental health phenomena in everyday life (10). EMA measures symptoms prospectively and dynamically in their environmental context through smartphone-administered questionnaires multiple times a day (10). EMA studies of psychopathology have provided key clinical insights (11). For instance, EMA has demonstrated that psychiatric symptoms fluctuate over time (12), and that the dynamics of these fluctuations can predict relapse and recovery from psychiatric disorders, independent of the symptoms’ average presence or absence (12, 13). Moreover, EMA provides an objective estimate of the dynamic interdependencies linking psychiatric symptoms to each other and their environmental context (14, 15). For instance, vulnerability to psychosis might be linked to short-term affective reactions to minor daily stressors (16). Conceptually, EMA findings support the view of psychiatric disorders as complex systems—networks of dynamic interactions between symptoms and contextual factors (17). Shedding light on these psychological and contextual dynamics may enhance our understanding and management of the heterogeneous clinical trajectories characterizing the emergence and recovery from psychiatric disorders (6, 18). While the rich clinical characterization provided by EMA has substantial potential, it also presents challenges for clinical translation (19). Psychiatric assessments must balance fidelity and accuracy with the intuitiveness needed for drawing reliable clinical conclusions (18, 20–23). Traditionally, psychometric instruments are designed according to the low-dimensional structure of correlation between items, in order to measure a limited number of latent phenomena that efficiently differentiate subjects from each other (24). However, this “psychometric method” does not apply to EMA for two reasons. First, from a statistical perspective, traditional dimensionality reduction techniques are not readily applicable to temporally structured EMA data (11), limiting our understanding of the low-dimensional structure of dynamic fluctuations in transient psychological states (8, 11, 25). Second, psychometric theory conceptually assumes that correlations between variables are epiphenomena of a limited number of underlying causal factors (24). This assumption justifies merging multiple variables or symptoms into dimensions or diagnostic labels that reflect such latent underlying mechanisms (26). However, evidence suggests that the latent structure of mental health phenomena could actually emerge from dynamic causal interactions between specific psychological and contextual factors (27). Clinically, this implies that capturing the true nature of mental health phenomena may be impossible if multiple symptoms are merged together (28) or if the intensity of inherently dynamic psychological/contextual phenomena is averaged over time (12, 13). EMA can measure the dynamic fluctuations of multiple psychological and contextual variables (10). However, it remains unclear how to best translate this wealth of information into a comprehensible and replicable view of the network structure of mental health phenomena (9, 29). In this paper, we introduce a novel framework for analyzing EMA data, aiming to overcome these challenges and unlock EMA’s full clinical potential. Specifically, we present Behavioral-Tractography, a method conceptually inspired by brain connectivity analysis (30, 31) and phenomenological descriptions of clinical reasoning (32). Behavioral-Tractography builds upon previously proposed Network-Analysis approaches to EMA (11, 33), which assess clinical patterns at their most granular level (33) by estimating the propensity of individual symptoms to co-occur using Mixed-Model Linear Regression (34). Graph theory algorithms then clarify the role of individual symptoms within psychological networks (31). While Network-Analysis ‘s symptom-level precision is valuable, it typically precludes a broader, low-dimensional view of large-scale clinical patterns (35). In the context of clinical reasoning, such an aggregated low-dimensional view is a crucial prerequisite for subsequent higher-resolution analyses of the roles of individual symptoms, akin to discerning the outline of the forest before focusing on individual trees (32). Thus, the first key innovation of Behavioral-Tractography is applying Network Dimensionality Reduction (NDR) techniques to EMA networks (36), which identified two highly consistent large-scale dimensions structuring symptom fluctuations in everyday life. By embedding high-dimensional symptom-level interactions within these large-scale coordinates, Behavioral-Tractography provides an innovative multi-scale view of clinical patterns that aligns conceptually with the clinical reasoning process (35). The second methodological innovation of Behavioral-Tractography lies in its mathematical and graphical representation of the temporal flow linking past and future behavioral variables. While cross-sectional variable associations are represented along the X-Y axis derived from NDR, time-lagged associations (computed using MMLR) are mapped along the Z-axis, linking past and future temporal layers. This novel 3D representation is both graphically intuitive and analytically advantageous. Inspired by brain connectivity analysis (30), Behavioral-Tractography enables the identification of bundles of pathways connecting past and future states along similar psychological-temporal trajectories. These 3D-trajectory bundles provide a novel “meso-scale” description of dynamic psychological interactions unfolding over time, which can be quantitatively compared across clinical populations.

We tested the Behavioral-Tractography framework by first validating its consistency across two independent cohorts of typically developing controls (TDs). We then explored its potential in the context of a complex psychiatric differential diagnosis—psychosis vulnerability (37) and Autism Spectrum Disorder (ASD) (38). Psychosis and ASD share multiple clinical features, including prominent social impairments (39, 40) and motivational difficulties (41–43). The challenge of differentiating these conditions (40, 44, 45) is still recognized in the DSM-5 diagnostic criteria for schizophrenia (46) and historically contributed to their misclassification as a single disorder (47, 48). However, accumulating evidence suggests that ASD and psychosis involve distinct or even opposing alterations in the dynamic interactions between psychological states and contextual factors (49–53). This divergence is thought to be driven by differences in the relative weight assigned to top-down expectations versus bottom-up sensory experiences of the environment (49–53).

We hypothesized that the Behavioral-Tractography framework could provide an intuitive and informative characterization of dynamic behavioral interactions that differentiate individuals with ASD from those at high genetic risk for psychosis due to 22q11.2 Deletion Syndrome (54), a well-validated, genetically homogeneous model of psychosis vulnerability (55–57). More broadly, we hypothesized that the Behavioral-Tractography framework could enhance the interpretability of EMA data and represent a step toward clinical translation. To facilitate its adoption, we developed a Behavioral-Tractography toolbox for analysis and visualization, which is freely accessible to the research community.

## Results

### Low-Dimensional Structure of Behavioral Fluctuations

We measured the cross-sectional association strength between each pair of EMA variables, using Linear Mixed Regression (LMR) (Figure-1-Panel-1). We analyzed the resulting symmetrical adjacency matrix, through Network Dimensionality Reduction (NDR) (36), to characterize the low-dimensional structure of behavioral fluctuations (Figure-1-Panel-2). We evaluated accuracy and consistency of NDR results across independent TD samples.

In both samples, only the first two NDR dimensions meaningfully represented network structure, as quantified by a strong negative correlation between Euclidean distance separating variables and the strength of their association (Table-2). Variable loadings on these dimensions were highly consistent across TD-Discovery and Replication samples (R-Dimension-1=0.95, p<0.0001; R-Dimension-2=0.93, p<0.0001, Figure-2-Panels-E1-E2). Additionally, NDR from the TD-discovery sample significantly predicted association strength between variables in the TD-replication sample (R=-0.54, p<0.0001, Figure-2-Panel-F). Notably, the predictive accuracy of training NDR directly on the replication sample was only marginally higher (R = −0.59, p < 0.0001) and not statistically different from NDR computed in the completely independent discovery sample (Fisher’s R-to-Z=-0.69, p=0.48, Figure-1-Panel-F1). These findings suggest the existence of a highly conserved two-dimensional structure underlying the coherent fluctuation of transient behavioral states. Nodes were positioned in a 2D space based on NDR loadings (Figure-2-Panels-A). We characterized such dimensions by examining clustering of behavioral variables along 4 network quadrants they defined.

– The Upper-right quadrant included variables such as *Sadness, Irritation, Anxiety* which define different forms of Affective-Distress.
– The Bottom-right quadrant included reversely coded psychological-wellbeing variables such as *Lacking-Happiness, Lacking-Relaxation* and *Lacking-Confidence* which described Lacking-Affective-Wellbeing.
– The Lower-left quadrant included *Lacking-Concentration, Lacking-Motivation, Lacking-Activity-Enjoyment* variables which described Lacking-Cognitive-Wellbeing.
– The Upper-right quadrant included psychotic-like experiences such as *Hallucinations, Feeling-Unsafe, Confusing-Reality-with-Imagination* which we defined as Cognitive-Distress.

Hence inspired by the work of Thomson and Tamir (58), we defined the differentiation of affective states (right) vs. cognitive states (left) as a first Rationality Dimension, while the differentiation of psychological distress (top) vs. lack of psychological wellbeing (bottom) defined a second Valence Dimension. This mapping also captured the correlation structure of contextual factors such as social isolation (*Being-Alone*), which clustered in the lower-left Lacking-Cognitive-Wellbeing quadrant but was linked to upper-right Affective-Distress variables via the subjective experience of *Loneliness*.

### Dissecting dynamic behavioral pathways through Behavioral Tractography in independent TD samples

While Network Dimensionality Reduction (NDR) provided a 2D mapping of co-occurring behavioral variables, it did not capture dynamic time-lagged associations between variables measured at consecutive EMA assessments, which we estimated with LMR. This yielded a 40×40 adjacency matrix, where variables were considered both as baseline predictors and as predicted outcomes at the next assessment, which we represented as a 3-Dimensional Multilayer-Temporal-Network. (Figure-1-Step-3). Cross-sectional connections were organized in 2 temporal layers. Within each layer, nodes were positioned along NDR dimensions, capturing their tendency to co-occur. Time-lagged associations between past and future states, linked Temporal-Layer-1 (TL1) on the left to Temporal-Layer-2 on the right, along a Z temporal dimension. This provided a unique coherent but differentiated representation of both propensity for variables to fluctuate together, and of dynamic behavioral interactions unfolding across time. The 3D angle of longitudinal connections further distinguished stability of psychological states, represented by horizontal connections, from dynamic shifts between different states, shown as diagonal connections (Figure-1-Step-4, Figure-3). To promote interpretability, we provide network-specific links to a dedicated open-source 3D visualization platform (mlnetwork-diplab.ch), fully integrated with the analysis pipeline: https://github.com/andreaimparato/Behavioral-Tractography-Toolbox.

Such large-scale 3D network view still conserved high-dimensional information on interactions between specific behavioral variables (31). We employed Dynamic Behavioral Centrality (DBC)(35) to measure the importance of individual variables in mediating behavioral pathways, linking past to future states across temporal layers (Figure-1-Panel-4). DBC results of were highly consistent results across TD discovery and replication samples (R=0.6, p<0.001, Figure-4-Panel-11). *Feeling-Tired* and *Lacking-Happiness* were key mediators at TL1 and TL2. *Lacking-Motivation* acted as a gateway in TL1, while *Anxiety* and *Sadness* served as funnels in TL2 (Figure-3-Panel-A). This would suggest that insufficient psychological wellbeing plays a gateway role in influencing future states, while affective distress states mainly act as funnels that perpetuate the effects of previous TL1 experiences on future TL2 variables. To further characterize such behavioral pathways, we combined Graph-Theory and NDR, through a novel Behavioral-Tractography pipeline. Through k-means clustering we dissected bundles composed of behavioral pathways with similar 3D trajectories, reflecting their role in mediating qualitatively similar dynamic behavioral interactions. The average trajectory of such Behavioral-Bundles provided a quantitative description of the main types of dynamic behavioral interactions. This is conceptually similar to dissecting meso-scale architecture of brain connectivity, based on 3D direction of water diffusion along axonal bundles, through MRI tractography (30).

We observed high consistency across independent TD samples, both in the composition of pathways assigned to each behavioral bundle (Rand-Index=0.83, p<0.001, Supplementary Figure-2-panel-A) and the resulting average bundle trajectory (p<0.0001, R=0.82, p<0.0001, Figure-4-Panels-12-13). Behavioral Diffusion Analysis (BDA) further characterized the role of individual variables within each bundle, which is conceptually reminiscent to dissecting micro-scale architecture of specific white-matter voxels, through diffusion tensor MRI (30). BDA vectors were significantly correlated between TD samples (R=0.32, p<0.0001), with stronger consistency for more central variables that mediated at least one path in both samples (R=0.54, p<0.0001, Figure-4-Panel-14). Here we provide a brief description of behavioral bundles which is detailed in Figure-4.

Bundles 1-2 connected psychological distress states in the upper portion of the network across time.

**Bundle-1**: TL1-Affective-Distress → TL2-Affective-Distress, mediated by TL1/TL2-*Anxiety*.

**Bundle-2**: TL1-Cognitive-Distress → TL2-Affective-Distress, mediated by TL2-*Sensory-Issues*, TL2-*Lacking-Relaxation*.

Bundles 3-4 connected lack of psychological well-being states in the lower portion of the network across time.

**Bundle-3**: TL1-Lacking-Affective-Wellbeing → TL2-Lacking-Affective-Wellbeing, mediated by TL1-*Lacking-Happiness* and TL1-*Lacking Confidence*.

**Bundle-4**: TL1-Lacking-Affective-Wellbeing → TL2-Lacking-Cognitive-Wellbeing (LCWB), mediated by TL1-Lacking Happiness, *TL1-Feeling-Tired*, TL1/TL2-*Lacking Motivation*.

Bundles 5-7 connected lack of psychological well-being in the lower portion of the network to psychological distress in the upper portion.

**Bundle-5**: TL1-Lacking-Affective-Wellbeing → TL2-Cognitive-Distress, mediated by TL1-*Irritation*, TL1-*Feeling-Unsafe*.

**Bundle-6**: TL1-Lacking-Cognitive-Wellbeing → TL2-Affective-Distress, mediated by TL1-*Lacking-Happiness*, TL1-*Lacking-Motivation* and TL2-*Sadness*.

**Bundle 7:** TL1-Lacking-Cognitive-Wellbeing → TL2-Cognitive-Distress, mediated by TL1-Social Isolation (*Being-Alone*, *Loneliness*), TL1/TL2-*Feeling-Tired*, TL2-*Lacking-Happiness*.

Bundles 8-9 connected psychological distress in the lower portion of the network to lack of psychological well-being in the upper portion.

**Bundle 8**: TL1-Cognitive-Distress → TL2-Lacking-Affective-Wellbeing (LAWB), mediated by TL1-*Feeling-Unsafe*, TL2-*Anxiety*, TL2-*Sadness*.

**Bundle 9**: TL1-Cognitive-Distress → TL2-Lacking-Cognitive-Wellbeing mediated by TL1/TL2-*Feeling-Tired*, TL2-*Lacking-Happiness*.

These results highlight the interwoven nature of behavioral interactions, providing a novel multi-scale view of key transition pathways across states.

### 3-Dimensional view behavioral dynamics differentiating clinical samples

Having evaluated consistency of Behavioral-Tractography across TD Samples, we tested its potential in distinguishing between 22q11DS and ASD clinical samples. NDR dimensions mapping behavioral fluctuations in both clinical groups were strikingly similar to each other and to TD samples (Figure-2-panels-B-C; Table 2), suggesting that NDR could provide a common coordinate system for navigating population-specific behavioral interactions.

Indeed, despite similar low-dimensional structure, we observed significant differences in network connectivity, which we tested against a null distribution of differences observed across randomly permuted populations. The strength of 88 individual connections differed in ASD-vs-22q11DS (p<0.001), 85 in ASD-vs-TD (p=0.02), and 60 in TD-vs-22q11DS (p=0.03). ASD networks exhibited a broadly distributed strengthening of cross-sectional connections relative to 22q11DS (Figure-3-Panel-F). The only increased cross-sectional connections in 22q11DS linked psychotic-like Cognitive-Distress variables (Upper-Left quadrant) to each other and to Affective-Distress. Compared to TD, ASD-related increases in cross-sectional connections primarily affected Affective-Distress variables, as shown by the Connectivity-Dissimilarity-Index (CDI) for *Anxiety* (p<0.001) and *Feeling-Unsafe* (p<0.001), which were more tightly linked to other Affective-Distress states and *Sensory-Issues* (p<0.001) (Figure-3-Panel-E). Along the vertical valence dimension, *Sensory-Issues* were also more strongly connected to Lacking-Cognitive-Wellbeing (*Lacking-Enjoying-Activity*), while Affective-Distress variables were more closely linked to Lacking-Affective-Wellbeing (*Lacking-Relaxation*). These findings suggest that in ASD, psychological states fluctuate as coherent, all-or-nothing ensembles and are more closely tied to contextual factors, such as environmental sensory properties.

The 22q11DS network showed an opposite trend, characterized by a general weakening of cross-sectional correlations compared to both ASD and TD (Figure-3-Panel-D). When compared to TD, reductions clustered in the lower Lacking-Wellbeing regions, with weaker correlations between Lacking-Affective-Wellbeing (*Lacking-Happiness*) and Lacking-Cognitive-Wellbeing (Lacking-*Motivation, Lacking-Concentration, Lacking-Enjoying-Activity, Being-Alone*, and *Feeling-Tired*, all p<0.001). In contrast, 22q11DS exhibited stronger longitudinal connections particularly in diagonal connections linking distinct behavioral states over temporal layers. This suggests a higher temporal dependency in 22q11DS, which was not driven by the temporal inertia of individual variables but rather reflected a higher propensity to transitions between qualitatively different states from one assessment to the next.

### Clinical differences in behavioral pathways revealed by Behavioral Tractography

We examined whether Behavioral-Tractography could provide additional insights on behavioral dynamics differentiating ASD and 22q11DS samples. In both groups, TL1-*Lacking-Happiness* emerged as a key gateway variable, while TL2-*Anxiety* and TL2-*Sadness* served as key funnel variables, mirroring DBC findings in TD. (Figure-3-Panels-B-C). However, TL1-*Feeling-Rejected* was an additional gateway variable in 22q11DS, whereas TL1-*Lacking-Enjoying-Activity* was a gateway variable in ASD. Similarly, TL2-*Lacking-Relaxation* and TL2-*Lacking-Confidence* were funnel variables in 22q11DS, while TL2-*Lacking-Happiness* was unique to ASD (Figure-3-Panels-B-C).

The trajectories of 7 out of 9 Behavioral-Bundles were highly similar across ASD and 22q11DS networks (Figure-5). However, Bundle-7 and Bundle-9 showed significant differences compared to randomly permuted network comparisons (Bundle-7: 27 pathways, p=0.01; Bundle-9: 29 pathways, p<0.001). Here we provide a brief description of population specific bundle trajectories, which are detailed in Figure-5. Behavioral-Diffusion-Analysis (BDA) results are detailed in Figure-5-Panel-10 and in supplementary material.

Bundle-9 linked TL1-Cognitive Distress variables to TL2-Lacking-Cognitive-Wellbeing variable. The 22q11DS Bundle-9 trajectory (Figure-5-Panel-9A), revealed a direct link between TL1 affective and social distress and subsequent Lacking-Cognitive-Wellbeing. Specifically, TL1-*Feeling-Rejected* (p<0.001) and TL1-*Loneliness* (p=0.04), directly influenced TL2-*Lacking-Concentration* (p=0.01), TL2-*Lacking-Motivation* (p=0.01), TL2-*Lacking-Excitement* (p=0.01), TL2-*Lacking-Enjoying-Activity* (p=0.03), TL2-*Lacking-Physical-Activity* (p<0.001), and TL2-*Being-Alone* (p=0.04). In ASD (Figure-5-Panel-9B), the effect of TL1-Cognitive-Distress on TL2-Lacking-Cognitive-Wellbeing was instead mediated by TL2-Affective-Distress. Specifically, TL1-*Sadness* (p<0.001), TL1-*Finding-Activity-Difficult* (p<0.001), and TL1-*Loneliness* (p=0.04) predicted TL2-*Sadness* (p=0.01), which was in turn associated with TL2-*Lacking-Relaxation* (p=0.02) and TL2-*Lacking-Happiness* (p<0.001). These findings suggest that in 22q11DS, social rejection associated with Psychotic/Cognitive-Distress states directly impacts motivational/cognitive well-being and social isolation. In contrast, in ASD, social isolation and functional impairment influence psychological well-being indirectly through secondary affective states (Figure-5-Panel-10; Supplementary-Video-1).

Bundle-7 mediated the reverse relationship, linking TL1-Lacking-Cognitive-Wellbeing to TL2-Cognitive-Distress, which in ASD was characterized by a direct trajectory (Figure-5-Panel-7B). Specifically, in TL1-*Lacking-Enjoying-Activity* (p<0.001) and TL1-*Struggling-with-Activity* (p<0.001) directly influenced TL2-*Sensory-Issues* (p<0.001) and TL2-*Loneliness* (p=0.04), mediating the effects of TL1-*Lacking-Concentration* (p=0.04). The 22q11DS Bundle-7 trajectory (Figure-5-Panel-7A), was instead indirectly mediated by TL2-Lacking-Affective-Wellbeing, particularly TL2-*Lacking-Happiness* (p<0.001). These results suggest that in ASD, insufficiently enjoyable activities directly contribute to psychological and social distress, whereas in 22q11DS, Lacking-Cognitive-Wellbeing effects are mediated by secondary Lacking-Affective-Wellbeing.

Confirmatory analysis (Supplementary-analysis-3) demonstrated high consistency in Bundle-7 and Bundle-9 trajectory differences, across random partitions of 22q11DS and ASD populations, in discovery and replication sub-samples. Overall, these findings highlight distinct, qualitative differences in bi-directional dynamic pathways linking Cognitive-Distress and Lacking-Cognitive-Wellbeing in ASD and 22q11DS.

## Discussion

In this paper we introduced and validated Behavioral-Tractography, a novel approach aiming to improve interpretability of behavioral data collected through Ecological Momentary Assessment (EMA). We firstly validated Behavioral-Tractography across two independent samples of TD individuals and then explored its clinical potential in differentiating individuals at high genetic risk for psychosis due to 22q11DS from individuals with idiopathic ASD.

### The implications of two-dimensional maps of behavioral states

EMA provides a dynamic and ecologically-embedded view of psychological phenomena that could complement current psychiatric assessments (4). However, the quest for precision/personalized care requires a delicate balance between richness/fidelity of assessments and intuitiveness required to draw clinically reliable conclusions (18, 20, 21). Mental health research has pursued this balance by characterizing broad dimensions that underline psychological variations across populations, in traits typically considered stable within individuals (59). EMA studies have however increasingly shown that psychological phenomena considered as stable (11, 60), such as mood (61) and personality (62), are more accurately represented as aggregates of transient mental states that fluctuate in relation to internal and external contextual factors (8). We still have a limited understanding of the low-dimensional structure of transient psychological states, as traditional dimensionality reduction techniques are not directly applicable to dynamic EMA data (11). The resulting predominantly detail-oriented focus on specific symptoms currently limits both interpretability and ability to generalize EMA results across studies and clinical populations.

In this paper, we applied a novel Network-Dimensionality-Reduction (NDR) approach to characterize the low-dimensional structure of transient psychological states. We show that psychological fluctuations measured with EMA could be mapped across two dimensions, reflecting Cognitive-vs-Affective symptoms and Psychological Distress vs Lack of Psychological Wellbeing with remarkable consistency across independent samples. Indeed, psychological mapping derived in a first TD-discovery-sample predicted the structure of behavioral fluctuations with remarkably similar accuracy in a completely independent TD-replication-sample. While the underlying mechanisms remain currently unclear, the low-dimensional structure revealed by NDR aligns with a recent series of studies demonstrating that, when estimating the probability of psychological transitions in others, psychological states are mapped according to 3 main dimensions, capturing rationality (cognition vs affect), valence (positive vs negative) and social impact (high-arousal-social vs low-arousal-asocial) (58, 63). The authors interpreted such low-dimensional psychological mapping with regards to its evolutionary value in navigating social interactions (63). Our results show that the same valence and rationality dimensions also predict fluctuations in how psychological states are subjectively experienced. This would suggest that the mapping of mental states in others could be tightly linked or even emerge from the first-person subjective experience of psychological fluctuations (64).

The rationality and valence dimensions observed in TDs were also remarkably consistent in 22q11DS and ASD clinical samples. This suggests that low-dimensional psychological mapping could provide a sufficiently conserved coordinate system, that might help clinicians compare identify and population-specific behavioral dynamics. The consistency of such psychological mapping could also carry significant implications for understanding the phenomenology of psychiatric disturbances (8). For instance, items measuring psychotic-like experiences - such as *Confusing-Reality-with-Imagination*, *Hallucinations,* or *Feeling-Unsafe* were consistently mapped in the upper-left Cognitive-Distress quadrant. This is highly consistent with phenomenological descriptions of the psychotic symptoms as a “cognitive” attempt to make sense of internally or externally generated events that are experienced as unusual, bizarre, or unexpected (65). Importantly, such EMA items were previously validated against a gold-standard clinical assessment of psychotic symptom severity in 22q11DS (66). Their consistent psychological mapping across TD and clinical samples therefore supports the view that psychotic manifestations vary in their intensity across individuals and contexts but share similar underlying subjective phenomenology (67, 68).

The differential mapping of psychological distress and Lacking-wellbeing states along the vertical valence dimension is also noteworthy, confirming that wellbeing is not sufficiently defined by the absence of psychological distress (69). Wellbeing states were interestingly also differentiated along the cognitive-to-affective dimension, separating cognitive wellbeing states - such as *Lacking-Concentration*, *Lacking-Motivation* or *Lacking-Enjoyment*– from affective wellbeing states - such as *Lacking-Happiness* or *Lacking-Relaxation*. This aligns with the distinction of Hedonic versus Eudaimonic wellbeing, rooted in both ancient philosophy and contemporary neuroscience (70). Hedonic wellbeing is characterized by positively-valanced affective states, including in particular tranquility and happiness (70), while Eudaimonic wellbeing is mainly characterized by vitality, motivation, and striving towards self-determined objectives with competence, autonomy and relatedness (70). Such distinction might also have some relationship with the modern cognitive-neuroscience differentiation of liking versus wanting phases of reward processing (71, 72). Importantly, assessment of psychological wellbeing remains at best a peripheral part of classical psychiatric consultations (69). Our results suggest that Behavioral-Tractography and EMA could help characterize the differential role of specific forms of psychological wellbeing in the pathophysiology of psychiatric disorders.

### The value of behavioral dynamics in differentiating clinical samples

The propensity for symptoms to manifest together however still does not capture dynamic interactions between psychological and contextual factors unfolding across time, which might play an important role in the pathophysiology of psychiatric disorders (17, 27). To this end, Behavioral-Tractography builds upon the low-dimensional mapping of psychological fluctuations to derive a novel 3D view of both cross-sectional and longitudinal dynamic interactions between symptoms. Such integrated 3D perspective intuitively differentiated 22q11DS and ASD from TD according to opposite differences in the dynamics of how behavioral states manifest and interact in daily life. 22q11DS was characterized by increased temporal dependencies, suggestive of an inability to recover from previous behavioral states. In ASD, mental events were instead more internally correlated at a single moment in time (i.e. in an “all-or-nothing” pattern), and more volatile across time, which could suggest heightened susceptibility to volatile contextual factors. Of note, traditional diagnostic assessments have struggled to differentiate the two conditions providing inconsistent estimates of the prevalence of ASD in 22q11DS (73). Our results suggest that an integrated view of behavioral dynamics could provide novel insights on otherwise similar psychiatric disorders.

We propose that diverging behavioral dynamics could be related to Bayesian models of perception (74–76) that have gained traction in their descriptions of both psychosis (51, 68) and autism (49, 52, 53, 68). According to Bayesian theories, the differential susceptibility to contextual factors can be understood as the relative propensity to modify internal probabilistic models of the world based on novel bottom-up sensory information (74, 75). According to Bayesian models ASD individuals might rely more strongly on recent sensory information (49, 53, 77), which would prove advantageous in repetitive environments requiring high sensory precision (52), but might be detrimental in more volatile contexts with lower signal/noise ratio, such as social interactions (53). ASD individuals might also be more susceptible to minor fluctuations in their current environment, which are less buffered by more stable prior models of the world, leading to higher volatility in psychological states (53). This description aligns with the more volatile and context-dependent psychological fluctuations which we observed in the ASD sample. Bayesian theories interestingly conceptualize psychosis as an opposite tendency to place excessive weight on internally-generated models of the world, leading to a predisposition to form strong assumptions on limited information, or even to become completely detached from reality as it is empirically perceived through the senses (51, 68). Given that internal models of the world are inherently less volatile than the contextual information from which they are generated, individuals at risk for psychosis should experience reduced volatility in their psychological states, in particular when compared to ASD (50, 78). Consistent with this description, in 22q11DS, psychological phenomena had a higher propensity to influence each other across time, particularly compared to ASD. Indeed, in accordance with Bayesian theories, our results suggest that ASD and Psychosis might not necessarily differ in the “amount” of environmental susceptibility, but rather in the dynamics with which internal psychological states are influenced by volatile contextual factors (53, 68). Measuring such behavioral dynamics through Behavioral-Tractography might therefore help differentiate otherwise similar psychiatric disorders.

### A novel multi-scale characterization of behavioral interactions

Moving from a static to a dynamic view of psychopathology, however, carries an inherent risk of providing information that is ultimately too complex to inform clinical reasoning (18, 20, 21). In this perspective, it was proposed that clinical reasoning approaches complexity flexibly and progressively, beginning with a macro-analysis of the outline of the forest, before a second level micro-analysis focusing on individual trees (32, 79). Behavioral-Tractography appears uniquely suited to accommodate and inform such a multi-scale view of clinical phenomena. Indeed, Behavioral-Tractography builds upon a large-scale view of behavioral fluctuations to dissect a limited number of Behavioral-Bundles linking qualitatively different states across time. Such Behavioral-Bundles also conserved high-resolution information on the role of individual variables, which were highly replicable across independent TD samples. For instance, Bundles-1-3 consistently characterized the gateway role of insufficient happiness and motivation in predicting subsequent psychological distress, while TL2-*Sadness* perpetuated the effects of previous psychological distress in Bundle-4.

In ASD and 22q11DS clinical samples, the general architecture and trajectory of most Behavioral-Bundles were largely conserved. However, Behavioral-Tractography still proved sufficiently sensitive to dissect differences in behavioral dynamics, which specifically affected cognitive/eudemonic wellbeing. In ASD, the Bundle-7 trajectory identified a direct pathway linking TL1-Lacking-Cognitive-Wellbeing, including *Lacking-Enjoying-Activity* and *Struggling-with-Activity*, to subsequent TL2-Cognitive-Distress, including *Feeling-Unsafe* and *Sensory-Issues*. This suggests that in ASD, the Cognitive-Wellbeing experience of enjoyment and mastery plays a key role in preventing subsequent psychological and social distress. Indeed, first-person accounts show that in ASD engagement in special interests is associated with increased affective wellbeing (80) and represents a way to mitigate anxiety (81). Moreover, treatment strategies prioritizing cognitive (eudemonic) wellbeing in ASD, by exploiting highly motivating interests to promote personal development and social engagement have been shown to be highly effective in ASD (91,92).

In 22q11DS, Cognitive-Wellbeing variables were instead characterized by weaker cross-sectional connections with concurrent affective variables, and stronger longitudinal connections with previous behavioral states. In particular, Bundle-9 in 22q11DS dissected a direct pathway linking social rejection associated to TL1-Cognitive-Distress psychotic-like experiences to TL2-Lacking-Cognitive-Wellbeing (*Lacking-Motivation, Lacking-Concentration*), which in in turn contributed to social isolation and affective flattening. The dissociation of the motivational difficulties from concomitant affective states is in line with a series of studies showing that negative symptoms of psychosis, including lack of motivation and anhedonia, are highly prevalent in 22q11DS (82, 83) and are predominantly linked to alterations in reward-related processes, notably initiation and reward anticipation (84, 85) more-so than to depressive symptoms (86). The trajectory of Bundle-9 in 22q11DS is interestingly directly reminiscent of the concept of secondary negative symptoms, which refers to the idea that negative symptoms, namely reduced motivation, reduced expressiveness and social deficits, can result from previous positive symptoms (87). Still, the concept of secondary negative symptoms is typically employed to describe clinical phenomena occurring over the course of weeks or months. The rapid temporal dynamics with which secondary motivational deficits were triggered from previous psychotic-like experiences is a novel finding that may have neurobiological implications, for reconciling the proposed opposing hyper and hypodopaminergic underpinnings of positive and negative symptoms (88). Of note, alterations in the motivational system are also highly implicated in the pathophysiology of ASD (41, 89, 90), which is in line with the key role of Cognitive-Wellbeing in ASD. However, we observed mirror-opposite behavioral dynamics linking cognitive/motivation wellbeing and distress in ASD and 22q11DS, suggesting that a dynamic view of dopaminergic motivation/reward system (91) might be instrumental in dissecting the differential pathophysiology of motivational and social deficits in ASD and psychosis.

## Conclusions and Limitations

This work should be considered as a proof of concept of the potential of a novel Behavioral-Tractography approach for the analysis of EMA data. We describe a novel low-dimensional structure of behavioral fluctuations that could have significant phenomenological implications and could prove clinically valuable in supporting the reliable description and comparison of behavioral dynamics across populations. Indeed, Behavioral-Tractography provides a novel multi-scale view of clinical phenomena that could intuitively accommodate and inform the clinical reasoning process (32). Developing tools to standardize this process will be essential for balancing personalization and reproducibility in future psychiatric care (79, 92, 93). Our clinical results support the view that embracing the behavioral complexity of mental health phenomena could help clarify differential pathophysiological mechanisms (6), while also addressing the growing clinical dissatisfaction towards reductionist models of psychopathology (27, 94).

The present work should however be considered in light of some limitations. Firstly, while 22q11DS is a highly validated clinical (95–99) and neurodevelopmental (100–104) model of psychosis vulnerability, providing a uniquely homogenous genetic pathophysiology against which to validate our assessment technique, the syndrome also presents some specificities, including more significant cognitive impairment (105, 106). However, supplementary analyses revealed similar results even when excluding 22q11DS with intellectual disability. Secondly, we did not explicitly test the added value of our approach in the clinical setting (107). For instance, future studies could evaluate their utility in helping clinicians and patients reach a more precise and shared understanding of their problems (108, 109) that is a cornerstone upon which therapeutic improvements are constructed (110).

## Methods

### Sample

A total of 236 individuals (113 females), aged from 11 to 31 years (mean age = 19.87, SD = 4.5), participated in the study. Participants included 60 individuals with ASD (31 females, mean age = 17.7, SD = 4.9), recruited from clinical centers in Geneva and France, as well as through family associations. Additionally, 70 carriers of 22q11DS (30 females, mean age = 19.5, SD = 4.8) were recruited from the Swiss 22q11DS longitudinal cohort. The study also included 53 typically developing (TD) individuals (TD-Discovery Sample: 26 females, mean age = 18.5, SD = 3.7) from the Geneva community and siblings of 22q11DS carriers. A separate replication sample of 53 TD individuals (TD-Replication Sample: 26 females, mean age = 19.8, SD = 4.2) was recruited from psychology students at the University of Geneva. All participants and their caregivers provided written consent and had a sufficient command of French. The study was approved by the Swiss Ethics Committee (CCER) in Geneva. ASD participants had a confirmed clinical diagnosis based on the Autism Diagnostic Observation Schedule (ADOS) (111)), and either the Autism Diagnostic Interview-Revised (ADI-R) (112) or the Social Communication Questionnaire (SCQ; (113)). The 22q11DS group had a confirmed genetic diagnosis of the 22q11.2 microdeletion through standard genetic testing methods. Comorbid psychiatric disorders in ASD and 22q11DS participants were assessed using validated diagnostic tools, including the Diagnostic Interview for Children and Adolescents-Revised (DICA),(114)) Schedule for Affective Disorders and Schizophrenia for School-Age Children (K-SADS-PL DSM-5) (115)) for participants under 18 years old, and the Structured Clinical Interview for DSM-IV Axis I (SCID-I; (116))), or the Structured Clinical Interview for DSM-IV/DSM-V (SCID-I/SCID-5-CV) (117)). Exclusion criteria for TD individuals included premature birth, a first-degree relative with a developmental disorder (except for a de novo 22q11.2 deletion), and a history of psychiatric, neurological, or learning disorders. Demographic details are provided in Table 1. Groups did not differ significantly in age or gender. IQ levels were comparable between TD and ASD groups, but 22q11DS individuals had significantly lower IQs. Additional analyses, excluding 22q11DS individuals with a Full-Scale IQ <75 (N=30/70), confirmed that the main findings remained unchanged.

**Table 1:** Demographic table describing Gender, Age, Full-Scale IQ, Average number of EMA assessments per subject, and Average number of consecutive EMA assessments per subject in the 3 populations, and results of statistical comparisons across populations. Gender differences are tested with Chi-Square test. Other continuous variables are reported as Mean **^±^** Standard Deviation and statistical significance is tested with Two-Sample T-Tests.

|  | Healthy Controls<br>N=53 | Healthy Controls<br>Replication<br>N=53 | 22q11DS<br>N=70 | ASD<br>N=60 | P-Value<br>HC-ASD | P-Value<br>HC-22q11<br>DS | P-Value<br>ASD-22q11<br>DS | P-Value<br>TD-D<br>TD_R |
| --- | --- | --- | --- | --- | --- | --- | --- | --- |
| Gender<br>(M/F) | 27/26 | 27/26 | 30/40 | 31/29 | p=0.78 | p=0.50 | p=0.32 | p=1 |
| Age | 18.5±3.7 | 19.8± 4.2 | 19.5 ± 4.8 | 17.7±<br>4.9 | p=0.37 | p=0.21 | p=0.04 | p=0.09 |
| Mean<br>Number of<br>Assessments | 29.4 ±<br>8.8 | 27.8 ± 8.8 | 27.1±<br>11.2 | 30 ± 9.4 | p=0.73 | p=0.21 | p=0.11 | p=0.34 |
| Number of<br>Consecutive<br>Assessments | 21.90 ±<br>7.82 | 19.53 ±<br>7.82 | 20 ± 9.96 | 22.22 ±<br>8.64 | p=0.84 | p=0.25 | p=0.18 | 0.12 |
| Full Scale<br>IQ | 111.2 ±<br>11.9 | | 71.9 ±<br>15.6 | 107.7 ±<br>15.9 | p=0.20 | p<0.001 | P<0.001 | |

### EMA assessment

The EMA protocol was administered via the RealLife Exp smartphone application over six days as described in detail elsewhere (66). Participants received semi-random notifications eight times daily between 7:30 AM and 10 PM, with at least 30 minutes between alerts. Each notification triggered the same EMA questionnaire, containing 33 to 38 items depending on conditional branching. Here, we analyzed the only non-conditional pre-branching portion of the questionnaire, consisting of 20 items rated on a 7-point Likert scale. The full EMA questionnaire is available in the Supplementary Material. The questionnaire assessed psychological distress (*Sadness, Anxiety, Anger*) and wellbeing (*Happiness, Confidence, Excitement, Relaxation, Motivation*), with wellbeing items reverse-coded for analysis. It also assessed the presence of psychotic-like experiences (*Hallucinations, Confusing-Reality-with-Imagination, Feeling-Unsafe*). Additionally, participants reported contextual details such as social setting (*Being-Alone* vs with others) and activity (current task). They also rated their experiences of social context (*Feeling-Lonely, Feeling-Rejected)* and activity (*Enjoyment, Concentration, Struggling*).

No significant group differences were found in the number of completed assessments or in the number of consecutively completed assessments. See Table 1 for details.

### Construction and dimensionality reduction of behavioral networks

The full analysis pipeline is described in detail and freely available at https://github.com/andreaimparato/Behavioral-Tractography-Toolbox. The pipeline described below was conducted separately for each population (TD-D, TD-R, 22q11DS and ASD), and on a global merged sample of all participants. In all cases, clinical variables were normalized across subjects using z-scores. We estimated association strength between cross-sectional variables from the same EMA assessment as the slope coefficient of a Linear Mixed Regression (LMR) with a random intercept for each subject; formalized as follows: *Y _ij_*=*β*_0_ + *β*_1_ *X _ij_* +*u_j_* + *ε_ij_* where *Y _ij_* is the outcome variable for subject *j* at observation *i, X _ij_* is the predictor variable for subject *j* at observation *i, β*_0_ is the fixed intercept, *β*_1_ is the fixed slope, *u _j_* is the random intercept per subject, *ε_ij_* is the residual error term using the lme function in MATLAB (MATLAB Version: R2024a)g. We performed this LMR procedure for each pair of variables, and filtered for associations that were significant at p<0.05 after correction for multiple comparisons using Benjamini Hochberg procedure (118). This produced a symmetric adjacency matrix, capturing the relative propensity for variables to fluctuate together across temporal assessments. See Figure 1, Step 1.

**Figure 1:**
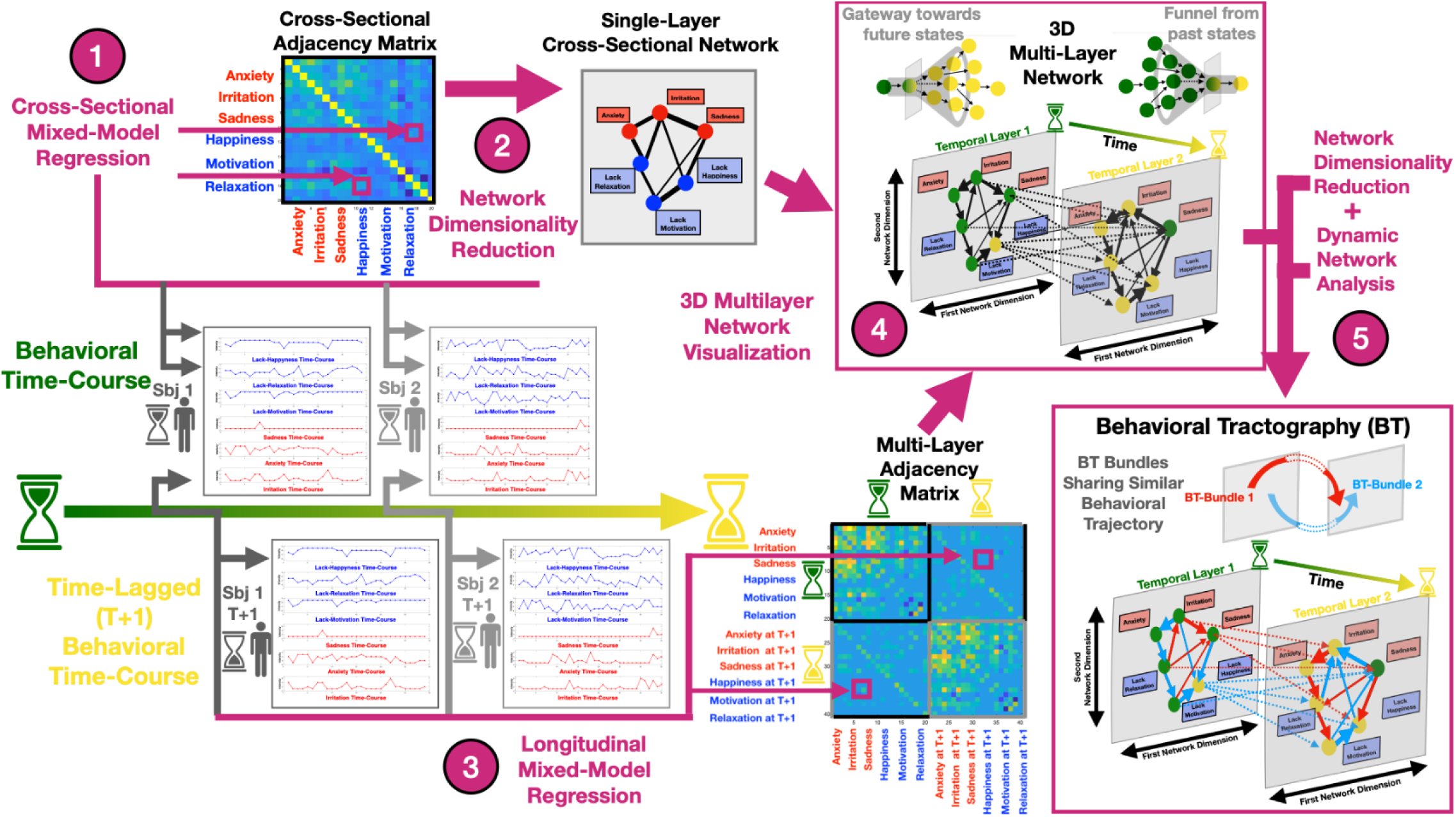
Schematic representation of methodological pipeline for Behavioral Tractography. The procedure is visually described in a simplified version considering behavioral time-courses of 6 variables in 2 subjects represented schematically in the center of the figure. **Step 1:** The strength of association between cross-sectional variables is estimated using Mixed Models Linear regression (MMLR), as the fixed slope coefficient, accounting for hierarchical data structure with a random intercept per subject. A symmetric cross-sectional adjacency matrix is constructed through iteration of MMLR for each pair of cross-sectional variables. **Step 2:** Nodes are positioned along X and Y axes according to network dimensionality reduction (NDR) performed on the cross-sectional adjacency matrix, grouping together variables that are more tightly associated. **Step 3:** The strength of longitudinal association between time-lagged variables is estimated using MMLR, as the fixed slope coefficient accounting for hierarchical data structure with a random intercept per subject. A Multi-Layer Adjacency Matrix is constructed through iteration of Longitudinal Mixed-Model Regression, between each pair of time-lagged variables. Each variable is considered twice, once as a baseline predictor at time T and once at the T+1 Time-Lagged assessment. Longitudinal associations are located in the upper right and lower left quadrants of the Adjacency matrix, while cross-sectional associations, computed in Step 1 are located in the upper-left and lower-right quadrants of the multi-layer adjacency matrix. **Step 4:** The Multilayer Network is represented in 3-Dimensional space. Cross-Sectional connections, computed in Step 1 are represented in two separate temporal layers. Within each temporal layer, nodes are positioned along X and Y axes according to network dimensionality reduction described in Step 2, capturing differential propensity for variables to co-occur at the same moment in time. Longitudinal time-lagged connections computed in Step 3 are represented as dashed lines along the Z axis, bridging across Temporal layer 1 (TL-1), located on the left, and Temporal Layer 2 (TL-2), located on the right. Dynamic network analyses applied to the Multi-Layer Adjacency Matrix are employed to identify shortest pathways connecting psychological contextual variables across temporal layers. The procedure identifies variables that play a disproportionate role in the dynamic evolution of behavioral variables across temporal layers. Such longitudinal hubs can be either identified in TL-1 and act as gateways towards future TL-2 variables, or in TL-2 and act as funnels that perpetuate the effects of previous TL-1 variables. **Step 5:** Behavioral-Tractography analysis dissects Bundles of pathways connecting psychological contextual variables across temporal layers with similar 3D Behavioral Trajectory. An average behavioral trajectory can be derived for each behavioral bundle by averaging the trajectory of pathways contained in it. Behavioral-Tractography retains information on the contribution of individual variables in each Behavioral Bundle.

**Figure 2.**
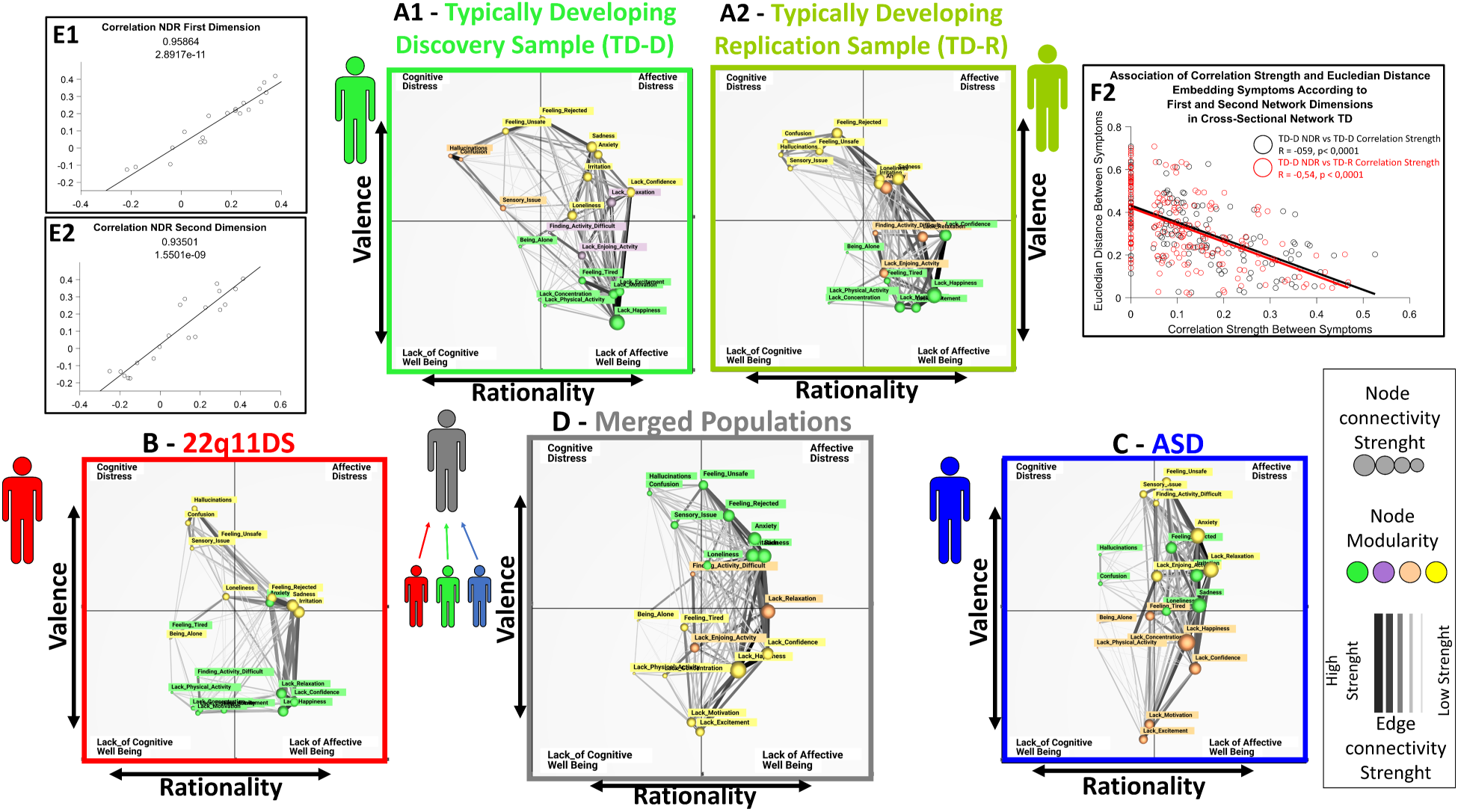
Single-layer networks representing cross-sectional patterns of co-occurrence between psychological and contextual variables at specific moments in time in different samples: **Panel A1:** TD Discovery Sample, **Panel A2:** TD Replication Sample, **Panel B:** 22q11DS, **Panel C**: ASD**, Panel D** Merged TD-D, 22q11DS and ASD samples. Each psychological and contextual variable is represented as an individual network node. **Thickness and transparency of network edges** represent the strength of cross-sectional statistical associations between variables. **Node size** represents the summed connectivity strength of the corresponding psychological or contextual variable. **Node color** is defined from modularity analysis and identifies subgroups of variables that are more tightly connected to each other that to the rest of the network. **Node position** is determined according to two main dimensions derived from network dimensionality reduction (NDR) and capturing the differential propensity for variables to manifest together. The first network dimension is represented horizontally and mainly captures rationality separating low-rationality affective states located on the right from high-rationality cognitive states located on the left. The second valence dimension is represented vertically, separating states of psychological distress located in the upper part of the graph from variables capturing lack of psychological well-being, in the lower part of the graph. These dimensions can be used to divide the network in 4 quadrants, populated by qualitatively distinct psychological states: Upper-Right-Quadrant: Affective Distress (AD); Lower-Right-Quadrant: Lack of Affective Well-Being (LAWB); Lower-Right-Quadrant: Lack of Cognitive Well-Being variables (L-CWB); Upper-Right-Quadrant: Cognitive Distress (CD). **Panels E1-E2:** Replication of NDR analysis across TD discovery and replication samples measured by Pearson correlation of loading of PC variables across NDR dimensions: E1: Replication of the first Rationality Dimension: E2: Replication of the second Valence Dimension. **Panel F: In Black:** Association of Euclidian Distance between Variables derived from NDR in TD Replication Sample and empirically measured correlation strength between variables in the same TD-Discovery sample, estimating overall accuracy of NDR representation. **In Red:** Association of Euclidian distance between variables derived from NDR in TD Discovery Sample and empirically measured correlation strength the sample TD-Replication sample, estimating the accuracy with which NDR can predict the structure of PC fluctuations measured in an independent TD sample.

Network Dimensionality Reduction (NDR)(36) was applied to uncover the low-dimensional structure of behavioral fluctuations. Specifically, we applied eigenvalue decomposition using MATLAB’s pca function to the above-mentioned cross-sectional adjacency matrix, as described in Sandini et al.(35). NDR dimensions that meaningfully characterize network structure group together variables that are more strongly associated, yielding a significant negative correlation between association strength and the Euclidean distance separating variables along NDR dimensions. In each population, we estimated accuracy of the first 3 NDR dimensions from such Euclidian-Distance/Association-Strength correlations. We also estimated consistency of NDR dimensions by correlating loadings of EMA variables across populations. See Table 2. Additionally, we estimated whether Euclidean distance defined according to NDR in the TD-Discovery sample, would predict association strength between variables in an independent TD-Replication sample. The first 2 NDR dimensions, which were the only ones to accurately characterize network structure in each population, were then used for two-dimensional mapping of cross-sectional networks (Figure 1, Step 2).

**Table 2:**
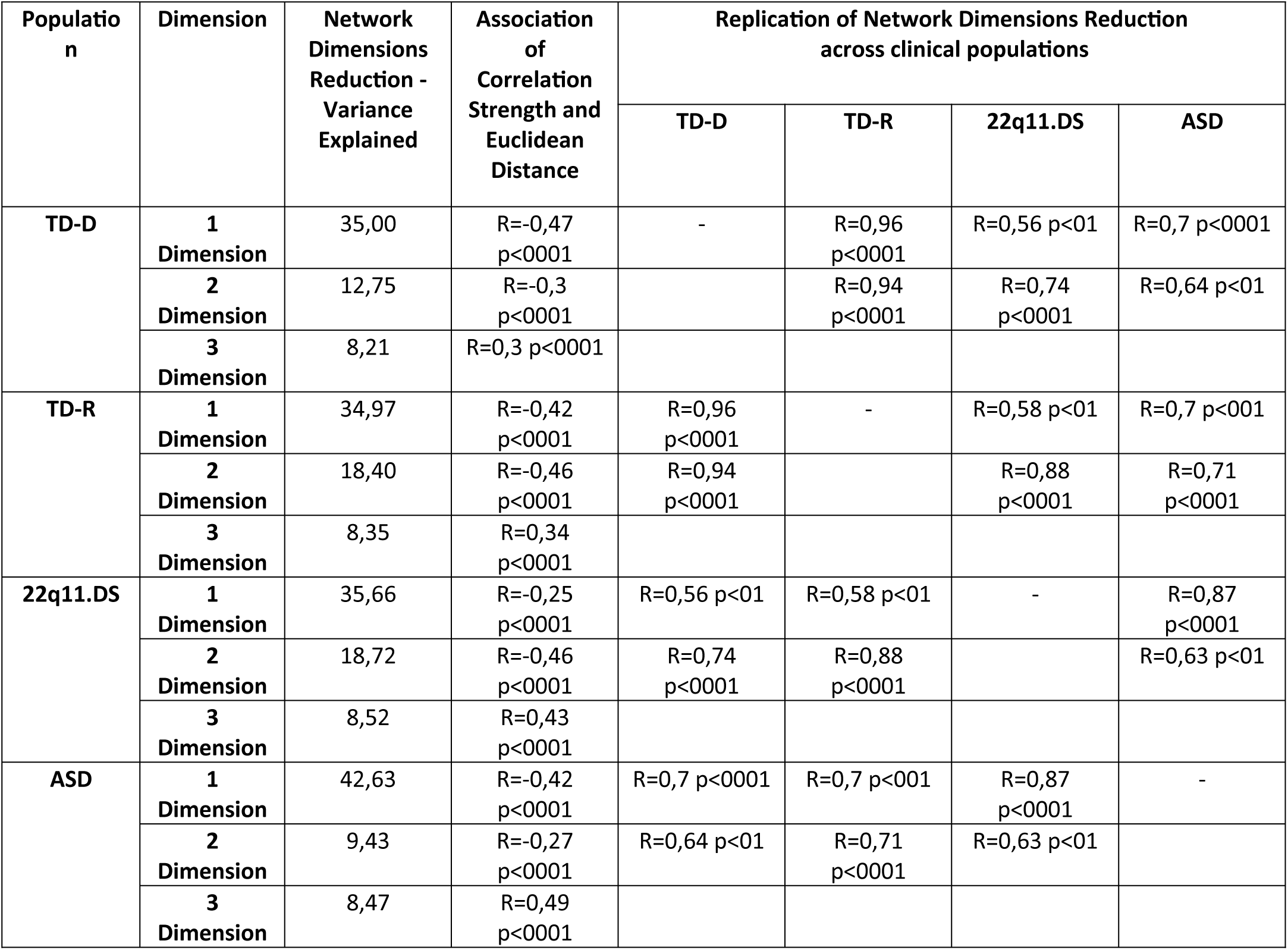
Description of Network Dimensionality Reduction (NDR) results in each samples and NDR replication across samples. The role of each of the first 3 network dimensions in accounting for network structure is described in terms of % of explained variance and in terms of correlation between Euclidian distance separating symptoms according to the specific dimensions and empirically observed correlation strength. A meaningful characterization of network architecture yields a significant negative correlation between Euclidian distance and correlation strength, implying that nodes that are represented closer to each other have a significantly higher tendency to fluctuate consistently. Consistency of NDR results across samples is measured by the Pearson correlation of loading of variables across the two main dimensions derived from NDR in each sample.

| Population | Dimension | Network Dimensions Reduction - Variance Explained | Association of Correlation Strength and Euclidean Distance | Replication of Network Dimensions Reduction across clinical populations |  |  |  |
| --- | --- | --- | --- | --- | --- | --- | --- |
|  |  |  |  | TD-D | TD-R | 22q11.DS | ASD |
| TD-D | 1 Dimension | 35,00 | R=-0,47<br>p<0001 | - | R=0,96<br>p<0001 | R=0,56 p<01 | R=0,7 p<0001 |
|  | 2 Dimension | 12,75 | R=-0,3<br>p<0001 |  | R=0,94<br>p<0001 | R=0,74<br>p<0001 | R=0,64 p<01 |
|  | 3 Dimension | 8,21 | R=0,3 p<0001 |  |  |  |  |
| TD-R | 1 Dimension | 34,97 | R=-0,42<br>p<0001 | R=0,96<br>p<0001 | - | R=0,58 p<01 | R=0,7 p<001 |
|  | 2 Dimension | 18,40 | R=-0,46<br>p<0001 | R=0,94<br>p<0001 |  | R=0,88<br>p<0001 | R=0,71<br>p<0001 |
|  | 3 Dimension | 8,35 | R=0,34<br>p<0001 |  |  |  |  |
| 22q11.DS | 1 Dimension | 35,66 | R=-0,25<br>p<0001 | R=0,56 p<01 | R=0,58 p<01 | - | R=0,87<br>p<0001 |
|  | 2 Dimension | 18,72 | R=-0,46<br>p<0001 | R=0,74<br>p<0001 | R=0,88<br>p<0001 |  | R=0,63 p<01 |
|  | 3 Dimension | 8,52 | R=0,43<br>p<0001 |  |  |  |  |
| ASD | 1 Dimension | 42,63 | R=-0,42<br>p<0001 | R=0,7 p<0001 | R=0,7 p<001 | R=0,87<br>p<0001 | - |
|  | 2 Dimension | 9,43 | R=-0,27<br>p<0001 | R=0,64 p<01 | R=0,71<br>p<0001 | R=0,63 p<01 |  |
|  | 3 Dimension | 8,47 | R=0,49<br>p<0001 |  |  |  |  |

### 3-Dimensional Network View of Psychological Contextual Dynamics

We examined whether the proposed low-dimensional structure of behavioral fluctuations could enhance the understanding of dynamic interactions between variables occurring from one EMA assessment to the next. Using the LMR procedure, we computed time-lagged associations between variables across consecutive assessments, excluding those interrupted by missing data or sleep. This resulted in a 40×40 adjacency matrix, where variables were represented both as baseline predictors and as predicted outcomes at the next assessment. Cross-sectional associations were mapped in the matrix’s upper left and lower right quadrants, while time-lagged associations occupied the off-diagonal quadrants (Figure 1, Step 3). To improve interpretability, we visualized behavioral dynamics in 3D. The time intervals between EMA assessments were represented along the Z-axis, with individual temporal layers displaying cross-sectional correlations. Within each layer, nodes were positioned on the X and Y axes according to their tendency to co-occur, as determined by NDR. Time-lagged associations were visualized as connections along the Z-axis, forming a 3D Multi-Layer Network.

This novel 3-Dimensional Multi-Layer Network provided a unique coherent but differentiated representation of both the cross-sectional propensity for variables to fluctuate together, and of dynamic interactions between variables unfolding across time. Moreover, the 3D angle of longitudinal connections effectively distinguishes stability of psychological states, represented by horizontal connections, from dynamic shifts between different states, shown as diagonal connections (Figure 1, Step 4; Figure 3 for a population-specific network). To enhance accessibility and facilitate clinical translation, we developed an open-source 3D network visualization platform (mlnetwork-diplab.ch). This tool integrates with MATLAB scripts for network construction and analysis and supports interactive 3D network exploration, including selective visualization of network components and is freely available at: https://github.com/andreaimparato/Behavioral-Tractography-Toolbox.

**Figure 3.**
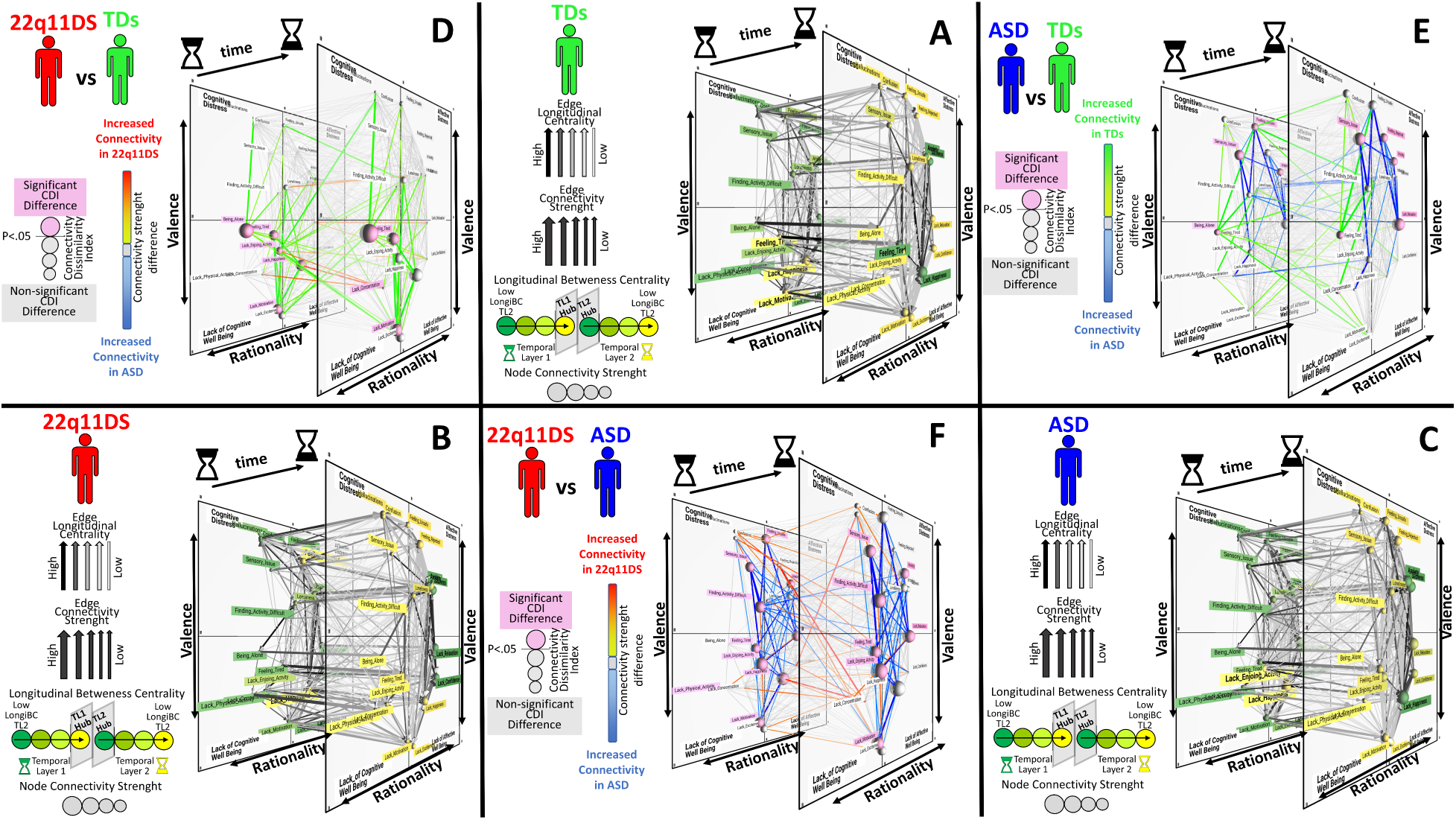
Differences in Multi-Layer Network structure across samples. **Panels A-B-C** depict Multi-Layer Temporal Networks constructed in the 3 populations separately; **Panel-A:** Healthy Control network, **Panel-B:** ASD network, **Panel-C:** 22q11DS network. **Cross-sectional connections** are represented within each temporal layer. **Longitudinal connections** are represented along the Z axis and bridge variables located in Temporal Layer 1 (TL-1) represented on the left to variables located in Temporal Layer 2 (TL-2) represented on the right. **Node position** within each temporal layer is determined according to the structure of cross-sectional networks as defined by network dimensionality reduction described elsewhere, grouping together variables that are more tightly related cross-sectionally. The slope of longitudinal edges reflects the propensity for dynamic interactions between qualitatively different behavioral states occurring across temporal assessments. **Thickness of network edges** represents the strength of statistical associations between variables derived from cross-sectional or longitudinal mixed-model-linear regression. **Arrows on connections** are derived from shortest path analysis applied to Multi-Layer Temporal Network (MLTN) and indicate that a particular edge, is part of a shortest path connecting variables across temporal layers. **Transparency of network** edges reflects the number of shortest paths that traverse through it. **Node color** reflects Longitudinal-Betweenness-Centrality. Green to Yellow color-coding is reverse coded in TL-1 and TL-2 so as to visually represent behavioral pathways flowing from Past States represented in green to future states represented in yellow. Specifically, TL-1 Variables are shaded from green to yellow according to their propensity to act as gateways towards future states in TL-2. TL-2 variables are shaded from yellow to green according to their propensity to act as funnels from past states in TL-1. Variables that act as significant longitudinal hubs are highlighted in bold. **Node size** represents the summed connectivity strength of the corresponding psychological or contextual variable. **Panels D-E-F** located in the sides of the triangular figure depict statistically significant differences in network structure across samples; **Panel-D:** Healthy Controls vs ASD, **Panel-E:** ASD vs 22q11DS **Panel-F** 22q11DS vs Healthy Controls. **Thickness and transparency of network edges** reflect the statistical significance of differences in cross-sectional and longitudinal association strength between variables across samples. **Color of edges** reflects the direction of the difference with green connections being increased in HCs, blue connections being increased in ASD and red connections being increased in 22q11DS. **Size of nodes** reflects the connectivity dissimilarity index (CDI), computed in the specific population contrast, with statistically significant differences highlighted in purple. **Node position** is determined with the same procedure described above. For each MLN figure we provide below a **link** to the MLNetwork platform for **dynamic online visualization**: **Panel A**: https://dev.mlnetwork-diplab.ch/3dvisualizer/net_hc/ **Panel B**: https://dev.mlnetwork-diplab.ch/3dvisualizer/22q_vs_hc/ **Panel C**: https://dev.mlnetwork-diplab.ch/3dvisualizer/22q_vs_asd/ **Panel D**: https://dev.mlnetwork-diplab.ch/3dvisualizer/hc_vs_asd/ **Panel E**: https://dev.mlnetwork-diplab.ch/3dvisualizer/22q_vs_hc/ **Panel F**: https://dev.mlnetwork-diplab.ch/3dvisualizer/22q_vs_asd/

### Multi-Scale dynamic view of behavioral interactions

The novelty of 3D-MLN lies in unique integration of a low-dimensional network structure with high-dimensional information on interactions between specific behavioral variables, which can be analyzed with the tools of Graph-Theory (31). The analysis pipeline outlined below is described in detail and available at: https://github.com/andreaimparato/Behavioral-Tractography-Toolbox.

We firstly used Floyd-Warshall shortest-path algorithm implemented in the Brain Connectivity Toolbox (31) to identify the shortest paths connecting EMA variables at past assessments represented in Temporal Layer one (TL1), to future variables at the next assessment, located in TL2. Each shortest-path was composed of single TL1èTL2 longitudinal edge and a variable number of TL1/TL2 cross-sectional edges. (Figure 5, Panel 8A). The number of shortest-paths traversing each network node, provides an estimate of the variables’ importance in mediating multiple dynamic behavioral interactions (35), which we define as Dynamic-Behavioral-Centrality (Dynamic-BC). Importantly, the Dynamic-BC of a given variable can differ significantly across Temporal Layers. Variables with high Dynamic-BC in TL1 may be considered as gateways, influencing multiple future states, while those in TL2 can be considered as funnels, that prevent recovery and perpetuate the effects of multiple previous psychological states (35). Statistically significant gateway and funnel variables were identified, by comparing the observed Dynamic-BC values against a null distribution derived from 1000 randomly constructed networks(35).

However, centrality measures alone do not capture the direction of pathway mediation, which may differ across populations. For instance, in Population-A Feeling-Rejected may mediate the effects of Anxiety on subsequent Social-isolation and Lacking-Happiness (**Population-A**: TL1-*Anxiety*==>TL1-*Feeling-Rejected*==>TL2-*Social-Isolation*==>TL2-*Lacking-Happiness*), while in Population-B Feeling-Reflected may play an equally important but different role mediating the effects of concomitant social isolation on subsequent Anxiety and Lacking-Happiness (**Population-B:** TL1-*Social-Isolation*==> TL1-*Feeling-Rejected*==>TL2-*Anxiety*==>TL2-*Lacking-Happiness*). See Figure 5, Panel 8A and 8B for an actual empirical representation of similar paths in two clinical populations. Measuring such mediation pathway direction may help generate hypotheses on differential underlying mechanisms, and potentially prove clinically useful in deciding to prioritize recognition and management of internal anxiety states, in Population A, or promote opportunities for social interaction in Population B.

Here, we combined Graph-Theory with NDR to provide a novel quantitative metric of such qualitative differences in behavioral pathways. We define this approach Behavioral-Tractography due to conceptual similarities with axonal tractography neuroimaging techniques to dissect axonal tracts based on water diffusion direction (30). In our 3D-Multi-Layer-Network framework, the XY position of network nodes defined by NDR, describes the main qualitative differences between co-occurring variables, while the left to right temporal axis differentiates simultaneous variables from those occurring in past and future assessments. The 3D trajectory of the paths linking network nodes hence provides a novel *quantitative* description of how behavioral variables interact dynamically across time. In the interest of interpretability, we firstly aimed at dissecting the meso-scale architecture of such behavioral trajectories, similar to the mesoscale architecture of bundles of axonal tracts (30). After having identified the shortest paths connecting behavioral variables across temporal layers, as described for Dynamic-BC, we decomposed each shortest-path trajectory in 4 sets of XYZ coordinates of its TL1-starting-node, TL1-exit-node, TL2-entry-node and TL2-ending-node. We then applied k-means clustering to these coordinates in order to dissect “bundles” of behavioral pathways that shared similar 3D trajectories, reflecting similar dynamic behavioral interactions. The optimal number of behavioral bundles was determined using a consensus procedure across all samples, described in supplementary material. See Figure 3, Panels 1-10, for a visual representation of Behavioral-Tractography results in the TD sample. We evaluated the consistency of Behavioral-Tractography results across independent TD-Discovery and TD-Replication samples, both in terms of composition of Behavioral-Tractography bundles derived from k-means clustering, and in terms of their average 3D trajectory.

We then complemented the meso-scale view of behavioral bundles with a micro-level characterization of the role of specific behavioral variables, which we define Behavioral-Diffusion-Analysis (BDA), due to conceptual similarities with Diffusion-Tensor-Imaging techniques to characterize micro-scale structural connectivity (119). Of note, such Diffusion-Tensor approaches can provide confounding results at the intersection of qualitatively different axonal tracts (119). Similarly, averaging the trajectory of all behavioral paths, would have yielded an incomplete or erroneous characterization of highly central variables, that mediate multiple qualitatively different behavioral interactions. To address this issue, we firstly dissected distinct behavioral bundles using Behavioral-Tractography and then averaged the XYZ coordinates of inputs and outputs for each variable in each bundle. This yielded average Behavioral-Diffusion vectors dissecting the *micro-scale* contribution of each variable to the “psychological movement” mediated by each *mesoscale* behavior bundle. We evaluated the consistency of BDA results across independent TD-Discovery and Replication samples. See Figure 3, Panel 10 for a visual representation of BDA analysis in the TD sample and supplementary material for a detailed description of each analysis step.

### Testing differences in 3D-Multi-Layer-Network measures across clinical samples

After evaluating their consistency across independent TD samples, we tested whether the proposed metrics could meaningfully dissect differences in behavioral dynamics in clinical samples. We tested differences in population specific networks through non-parametric permutation testing. For each comparison (ASD vs. 22q11DS, TD vs. ASD, 22q11DS vs. TD), subjects were randomly reassigned to equally sized groups for 1000 iterations, for which networks were constructed and compared. This provided a null distribution, against which to test network differences observed across populations. We first examined differences in the strength of individual network connections, correcting for family-wise errors by assessing the proportion of significant connections (p<0.05) in a null distribution of 1000 randomly separated subjects. We also measured the average difference in connectivity strength for each variable computing a Connectivity Dissimilarity Index (CDI) which we tested also against randomly permuted network comparisons.

Next, we tested differences in the trajectory Behavioral-Tractography bundles. Differences in bundle trajectory dissected in independent networks, can be influenced by both bundle composition (which TL1-Starting-Variables are connected to which TL2-Ending-Variables) and by the actual trajectory of pathways assigned to each bundle. In the interest of interpretably, we adapted the Behavioral-Tractography to selectively focus on the latter, through a coupled k-means clustering procedure, performed on pathway coordinates measured in both populations (Start-TL1, Exit-TL1-Population-A, Exit-TL1-Population-B, Entry-TL2-Population-A, Entry-TL2-Population-B, End-TL2). The resulting bundles connected a shared set of TL1 to TL2 variables across populations, while still allowing for potential population-specific differences in behavioral trajectories. (Figure 4, Panels 1-10). To quantify such differences, we computed the Euclidian distance between pathways coordinates across populations, which we divided by the average Euclidian distance separating population-specific pathways in each bundle, to normalize across-population differences for within-population consistency in bundle trajectory. We compared such values against a null distribution of randomly defined network comparisons.

**Figure 4.**
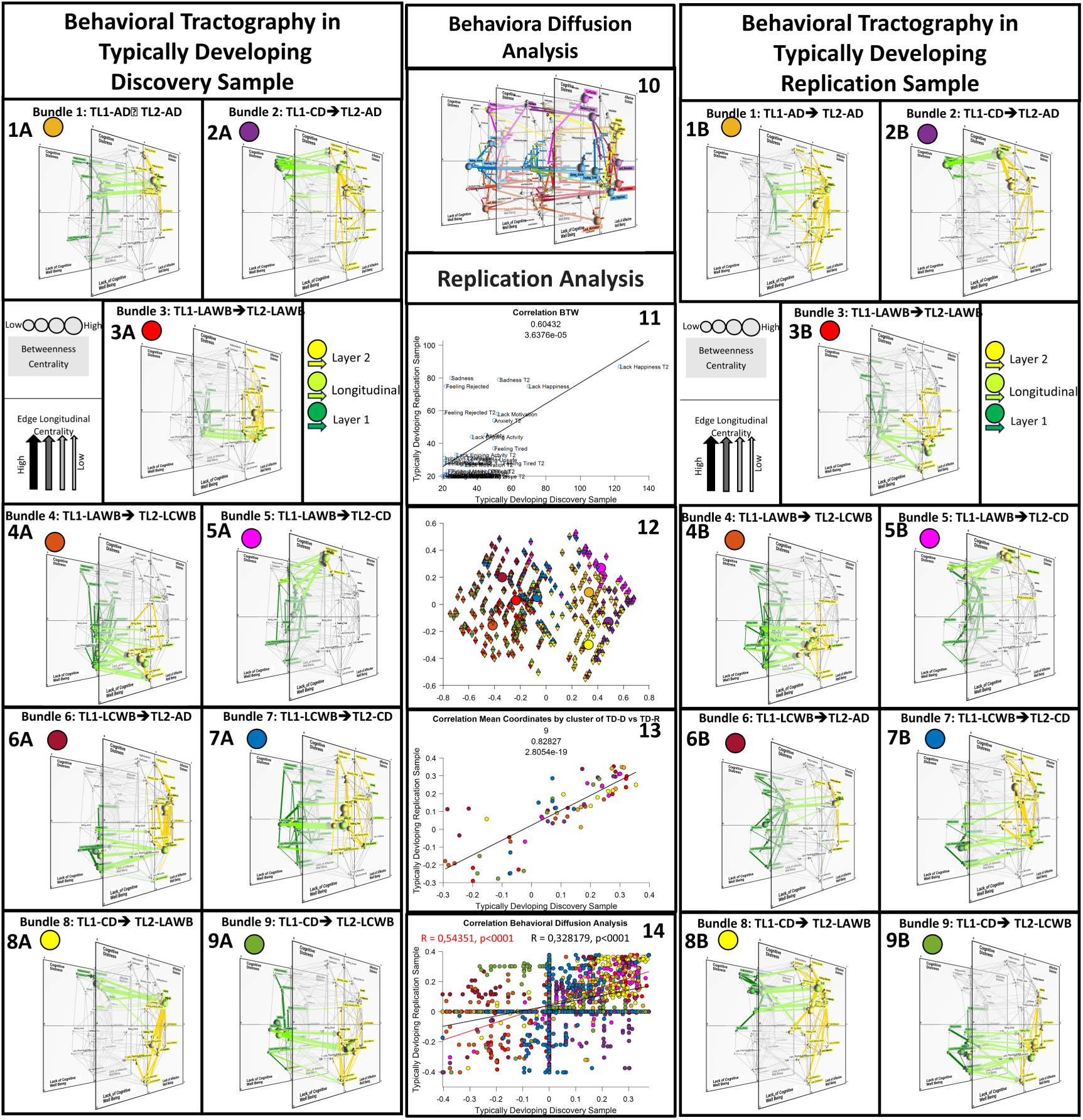
Results of Behavioral-Tractography and Behavioral Diffusion Analysis (BDA) in Typically Developing Discovery (TD-D) and Replication (TD-R) samples. **Panels 1A-1B:** Bundle 1: TL1-AD-->TL2-AD. **Panels 2A-2B:** Bundle 2: TL1-CD-->TL2-AD. **Panels 3A-3B:** Bundle 3: TL1-LAWB-->TL2-LAWB. **Panels 4A-4B:** Bundle 4: TL1-LAWB -->TL2-LCWB. **Panels 5A-5B:** Bundle 5: TL1-LAWB -->TL2-CD. **Panels 6A-6B**: Bundle 6: TL1-LCWB-->TL2-AD. **Panels 7A-7B:** Bundle 7: TL1-LCWB -->TL2-CD. **Panels 8A-8B:** Bundle 8: TL1-CD-->TL2-LAWB. **Panels 9A-9B:** Bundle 9: TL1-CD-->TL2-LCWB. Green to Yellow color coding reflects the dynamic progression of PC paths from TL1 to TL2 (Start-TL1==>Exit-TL1: Dark Green; Exit-TL1==>Entry-TL2: Light Green; Entry-TL2==>End-TL2: Yellow) **Panels 10:** BDA trajectories of variables that disproportionately mediate PC pathways in each Behavioral Bundle in TD-D Sample. Color coding of BDA vectors reflects the BT bundle that is significantly mediated by the corresponding variables. Variables that act as gateways for subsequent PC states are represented in TL-1 with their respective BDA trajectories connecting TL-1 and TL-2. Variables that act as funnels for previous PC states are represented in TL-3 with their respective BDA trajectories connecting TL-2 and TL-3. **Panel 11:** Replication analysis of Dynamic Betweenness Centrality across TD-D and TD-R samples. **Panel 12:** Replication analysis of Behavioral Tractography across TD-D and TD-R samples in terms of composition of Behavioral Bundles. Each triangle represents a Behavioral pathway connecting variables across temporal layers. Pathways are plotted in two-dimensional space derived from Principal-Component-Analysis of their 3D trajectory. PCA was performed on the same data-matrix to which K-means was applied to dissect behavioral bundles, which was composed of a line for each pathway each and 8 columns codifying it’s 3-D Trajectory (X-Y-Starting-TL1, X-Y-Exit-TL1, X-Y-Entry-TL2, X-Y-End-TL2). Color of triangles represent the attribution of pathways to one of 9 Bundles. Upward-pointing triangles represent composing Behavioral-Bundles in TD-D sample and downward-pointing triangles in TD-R sample. **Panel 13:** Replication of average 3D coordinates of the 9 Bundles computed in independent TD-D and TD-R sample. Each point represents one of 8 coordinates for each bundle. **Panel 14:** Replication analysis of BDA trajectories computed for each variable and each bundle across TD-D and TD-R samples. BDA trajectories are codified by 8 XY coordinates and computed across 9 clusters for all 40 network nodes and yielding a total of 2880 points that are correlated across samples, as represented by the black regression line. When a given variable didn’t mediate any PC-path in a given bundle the corresponding BDA coordinates were set to 0. We also repeated the BDA replication analysis considering only variables that mediated at least one path in the corresponding Behavioral-Bundle in both samples, as represented by the red regression line. For each MLN figure we provide below a **link** to the MLNetwork platform for **dynamic online visualization**: **Panel 1A**: https://dev.mlnetwork-diplab.ch/3dvisualizer/cl4_tdds/ **Panel 1B**: https://dev.mlnetwork-diplab.ch/3dvisualizer/cl4_tdrs/ **Panel 2A**: https://dev.mlnetwork-diplab.ch/3dvisualizer/cl7_tdds/ **Panel 2B**: https://dev.mlnetwork-diplab.ch/3dvisualizer/cl7_tdrs/ **Panel 3A**: https://dev.mlnetwork-diplab.ch/3dvisualizer/cl1_tdds/ **Panel 3B**: https://dev.mlnetwork-diplab.ch/3dvisualizer/cl1_tdrs/ **Panel 4A**: https://dev.mlnetwork-diplab.ch/3dvisualizer/cl3_tdds/ **Panel 4B**: https://dev.mlnetwork-diplab.ch/3dvisualizer/cl3_tdrs/ **Panel 5A**: https://dev.mlnetwork-diplab.ch/3dvisualizer/cl6_tdds/ **Panel 5B**: https://dev.mlnetwork-diplab.ch/3dvisualizer/cl6_tdds/ **Panel 6A**: https://dev.mlnetwork-diplab.ch/3dvisualizer/cl2_tdds/ **Panel 6B**: https://dev.mlnetwork-diplab.ch/3dvisualizer/cl2_tdrs/ **Panel 7A**: https://dev.mlnetwork-diplab.ch/3dvisualizer/cl9_tdds/ **Panel 7B**: https://dev.mlnetwork-diplab.ch/3dvisualizer/cl9_tdrs/ **Panel 8A**: https://dev.mlnetwork-diplab.ch/3dvisualizer/cl5_tdds/ **Panel 8B**: https://dev.mlnetwork-diplab.ch/3dvisualizer/cl5_tdrs/ **Panel 9A**: https://dev.mlnetwork-diplab.ch/3dvisualizer/cl8_tdds/ **Panel 9B**: https://dev.mlnetwork-diplab.ch/3dvisualizer/cl8_tdrs/ **Panel 10:** https://dev.mlnetwork-diplab.ch/3dvisualizer/btw_tdds/

**Figure 5.**
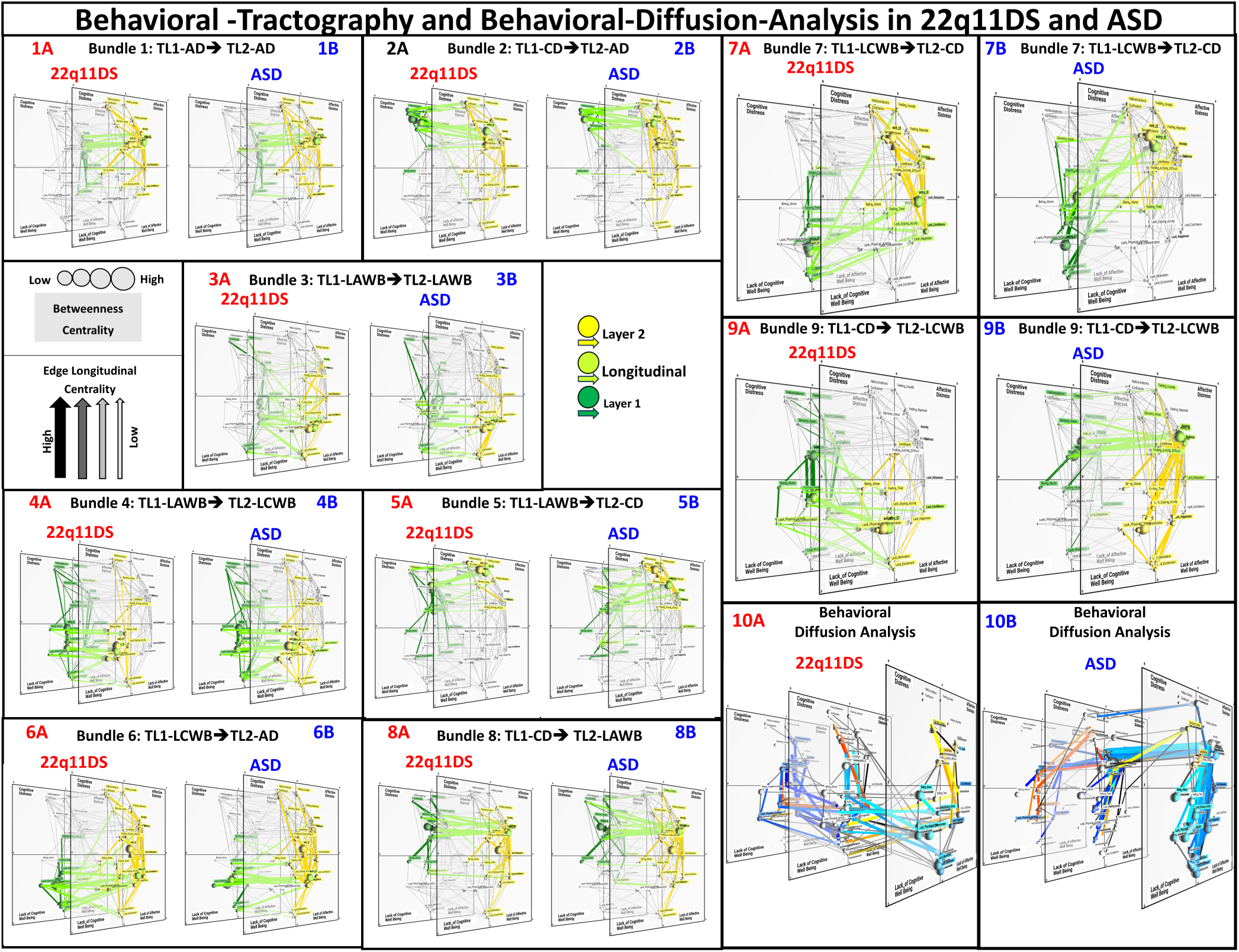
Population specific PC pathways in 22q11DS and ASD samples as described by Behavioral-Tractography and Behavioral Diffusion Analysis (BDA). **Panels 1-9:** Results of Behavioral-Tractography in 22q11DS **(A)** and ASD **(B);** Green to Yellow color coding reflects the dynamic progressing of PC paths from TL1 to TL2 (Start-TL1==>Exit-TL1: Dark Green; Exit-TL1==>Entry-TL2: Light Green; Entry-TL2==>End-TL2: Yellow) **Panels 1A-1B:** Bundle 1: TL1-AD-->TL2-AD. **Panels 2A-2B:** Bundle 2: TL1-CD-->TL2-AD. **Panels 3A-3B:** Bundle 3: TL1-LAWB-->TL2-LAWB. **Panels 4A-4B:** Bundle 4: TL1-LAWB -->TL2-LCWB. **Panels 5A-5B:** Bundle 5: TL1-LAWB -->TL2-CD. **Panels 6A-6B**: Bundle 6: TL1-LCWB-->TL2-AD. **Panels 7A-7B:** Bundle 7: TL1-LCWB -->TL2-CD. **Panels 8A-8B:** Bundle 8: TL1-CD-->TL2-LAWB. **Panels 9A-9B:** Bundle 9: TL1-CD-->TL2-LCWB. **Panels 10A-10B:** Results of BDA for variables in 22q11DS (Panels 10A) and ASD (Panels 10B). Warm colors identify variables that drive significant differences in Bundle-9 (TL1-LCWB==>TL2-CD). Cold colors identify variables that drive significant differences in Bundle-7 (TL1-CD==>TL2-LCWB). BDA trajectories reflecting differential functional role of TL1-Variables connect TL1 to TL2, while BDA trajectories reflecting differential functional role of TL2-Variables connected TL2 to TL3. The individual PC paths that contribute to average BDA trajectories are represented in Black. For each MLN figure we provide below a **link** to the MLNetwork platform for **dynamic online visualization**: **Panel 1A**: https://dev.mlnetwork-diplab.ch/3dvisualizer/cl4_22q/ **Panel 1B**: https://dev.mlnetwork-diplab.ch/3dvisualizer/cl4_asd/ **Panel 2A**: https://dev.mlnetwork-diplab.ch/3dvisualizer/cl7_22q/ **Panel 2B**: https://dev.mlnetwork-diplab.ch/3dvisualizer/cl7_asd/ **Panel 3A**: https://dev.mlnetwork-diplab.ch/3dvisualizer/cl1_22q/ **Panel 3B**: https://dev.mlnetwork-diplab.ch/3dvisualizer/cl1_asd/ **Panel 4A**: https://dev.mlnetwork-diplab.ch/3dvisualizer/cl3_22q/ **Panel 4B**: https://dev.mlnetwork-diplab.ch/3dvisualizer/cl3_asd/ **Panel 5A**: https://dev.mlnetwork-diplab.ch/3dvisualizer/cl6_22q/ **Panel 5B**: https://dev.mlnetwork-diplab.ch/3dvisualizer/cl6_asd/ **Panel 6A**: https://dev.mlnetwork-diplab.ch/3dvisualizer/cl2_22q/ **Panel 6B**: https://dev.mlnetwork-diplab.ch/3dvisualizer/cl2_asd/ **Panel 7A**: https://dev.mlnetwork-diplab.ch/3dvisualizer/cl9_22q/ **Panel 7B**: https://dev.mlnetwork-diplab.ch/3dvisualizer/cl9_asd/ **Panel 8A**: https://dev.mlnetwork-diplab.ch/3dvisualizer/cl5_22q/ **Panel 8B**: https://dev.mlnetwork-diplab.ch/3dvisualizer/cl5_asd/ **Panel 9A**: https://dev.mlnetwork-diplab.ch/3dvisualizer/cl8_22q/ **Panel 9B**: https://dev.mlnetwork-diplab.ch/3dvisualizer/cl8_asd/ **anel 10A**: https://dev.mlnetwork-diplab.ch/3dvisualizer/bda_22q/ **Panel 10B**: https://dev.mlnetwork-diplab.ch/3dvisualizer/bda_asd/

As a further confirmatory analysis, we randomly sub-divided 22q11DS and ASD populations in equally sized independent discovery and replication subsamples, for 500 iterations. We estimated both consistency of group differences in bundle trajectory across bootstrapped discovery samples, and their replicability across independent discovery and replication samples, comparing such values against a null-distribution of randomly assigned diagnostic labels. A detailed description of this analysis is provided in supplementary material (Supplementary-analysis-3). Finally, Behavioral-Diffusion-Analysis was applied to determine the role of specific variables in mediating differences within behavioral bundles that significantly varied across samples. Results of BDA comparisons between 22q11DS and ASD samples are shown in Figure 4, Panel 10 and detailed in the following sections.

## Data Availability

All data produced are available online at https://github.com/andreaimparato/Behavioral-Tractography-Toolbox

## Acknowledgments

This study was supported by the Swiss National Science Foundation (SNSF) (Grant numbers: to SE FNS 320030_179404, FNS 324730_144260) and by the National Center of Competence in Research (NCCR) Synapsy-The Synaptic Bases of Mental Diseases (SNF, Grant number: 51AU40_125759). Prof Maude Schneider (#162006) and Dr Corrado Sandini (#209096) were supported by grants from the SNF. We thank Charlotte Dubois, Lou Mattei, Célia Charbonnier, Sonia d’Emma, Kilian Bruttin, Sara Copetti et Angélique Bugnon for their help with data collection. We warmly thank all the families who participated in the study.

