## Supplementary_Material for "More than the sum of their parts: A Behavioral Tractography analysis of distress wellbeing and context reveals dynamic signatures of psychosis and autism"

**Supplementary Analysis 1: Selection of the optimal number of clusters for Behavioral Tractography Analysis**

**Methods**

The optimal number of clusters was initially defined in each population separately. To this end, we employed the Gap algorithm implemented in the GapEvaluation MATLAB function (https://ch.mathworks.com/help/stats/clustering.evaluation.gapevaluation.html), requesting evaluations for cluster counts ranging from 2 to 10. To address the potential ambiguity inherent in K-means clustering, due to the initial centroid selection, we applied the Gap criterion to 1000 versions of the K-means algorithm, varying the randomization used for initial centroid selection at each iteration. We utilized the k-means++ algorithm as the default method for initial centroid selection in MATLAB's K-means implementation. This approach yielded a distribution of optimal K values, accounting for the non-deterministic nature of k selection and the impact of the initial centroid choice on the final output. Subsequently, we compared the results obtained for each population. To facilitate interpretability of differences in network structure across populations, which was the essential aim of the Behavioral-Tractography procedure, we defined a single optimal number of clusters across all four populations, which was defined as the median of optimal K values computed in the 4 populations.

**Results**

Our analysis revealed distinct optimal cluster numbers for each population: 8 clusters for individuals 22q11DS, 9 clusters for ASD, 10 clusters for TD-D, and 9 clusters for TD-R, which yielded a median value of 9 clusters across populations. The distribution of optimal K values computed across 1000 iterations in each population is displayed in Supplementary Figure 1.

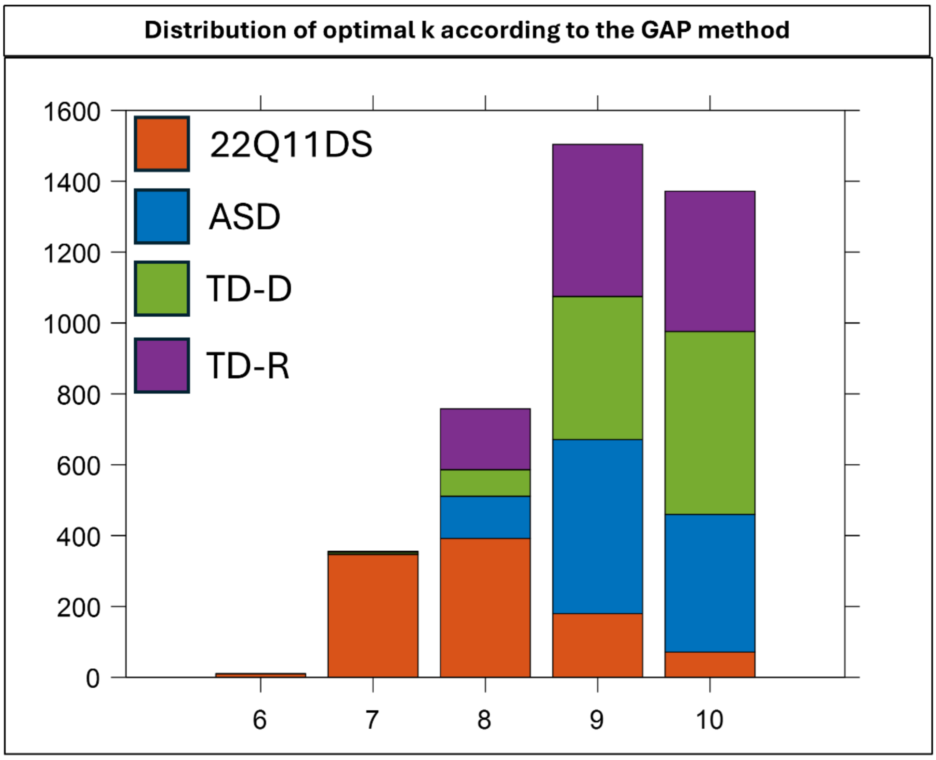


**Supplementary figure 1**: Distribution of optimal K values according to Gap criterion computed in each population across 1000 iterations of the K-means algorithm, varying the initial distribution of centroids at each iteration.

**Supplementary Analysis 2: Replication of Behavioral Tractography across Typically Developing Discovery and Replication Samples.**

In this analysis, we evaluated the replicability of the K-means clustering procedure employed to define Behavioral-Tractography bundles across Typically Developing Discovery and Replication samples (TD-D and TD-R). We firstly estimated the similarity of K-means clustering results, in terms of the extent to which paths were assigned to the same clusters in the TD-D and TD-R samples. To this end we employed the Rand Index (RI), a widely used measure to assess the similarity between two sets of clustering results. The RI quantifies the level of agreement between the cluster assignments by evaluating the proportion of pairwise agreements and disagreements with. RI values can range from 0 to 1, where 1 indicates perfect concordance in clustering results.
We computed the RI between TD-D and TD-R, which we then compared against a null distribution obtained by permuting the TD-R clustering 1000 times, Supplementary Figure 2 Panel A.
We also evaluated whether the correct attribution of paths to a given cluster across the two populations was moderated by the Euclidian distance between the path and the centroid of the corresponding cluster, which reflects the extent to which the trajectory of a given path is reflected in by the average trajectory of the corresponding Behavioral-Tractography bundle. We tested whether paths that more tightly reflected the average trajectory of the Behavioral-Tractography bundle had a higher likelihood to be consistently assigned to the same cluster across populations, which would further validate the consistency of Behavioral-Tractography results across populations, Supplementary Figure 2 Panel B.
After assessing the replicability of the composition of paths attributed to each Behavioral-Tractography cluster, we evaluated the qualitative similarity in the 3D trajectories of the resulting Behavioral-Tractography bundles. As described in the main text, the trajectory of each Behavioral-Tractography bundle was codified by the X and Y coordinates of the TL1-Starting-Point, TL1-Exit-Point, TL2-Entry-Point and TL2-End-Points. We correlated then means of XY coordinates across populations to estimate the qualitative similarity in the 3D trajectory of Behavioral-Tractography bundles, Supplementary Figure 2 Panel C.

**Results**

Overall, the K-means clustering procedure yielded highly consistent results with a Rand Index of 0.832 that was significantly higher than would be expected by chance $p<0.001$ (See Supplementary Figure 2, Panel A). Moreover, paths that had inconsistent cluster assignment across populations also had a significantly higher Eucledian distance from their corresponding cluster centroids ($p<0.001$; See Supplementary Figure 2, Panel B). This would indicate that paths that more strongly contribute to the average trajectory of the corresponding Behavioral-Tractography-Bundle had also a higher likelihood of being consistently assigned across samples, which further validates the replicability of our Behavioral-Tractography analysis across samples. Finally, the average trajectory of Behavioral-Tractography bundles was highly consistent across populations as indicated by the strong positive correlation of the average XY coordinates of Behavioral-Tractography bundles across populations ($R=0.82$, $p<0.001$; See Supplementary Figure 2, Panel C).

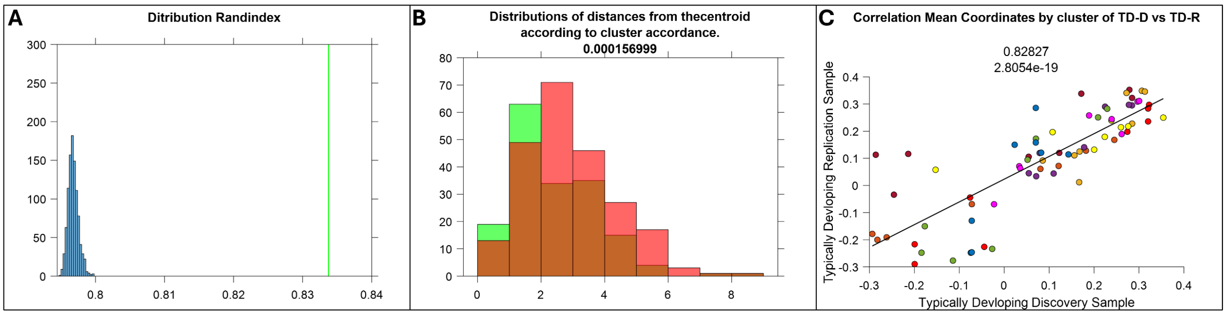


**Supplementary Figure 2:** Replication Behavioral Tractography Analysis across Typically Developing Discovery and Replication Samples.
**Panel A**: Empirically observed Rand Index value measuring the correspondence of K-means clustering results across TD-D and TD-R samples represented in green, compared against the null-distribution of the Randindex values computed across 1000 randomly permuted clustering solutions, represented in blue.
**Panel B**: Distribution of Euclidian distances between the trajectory of paths and the centroid of the cluster they were assigned to, for paths that were consistently assigned to the same cluster in TD-D and TD-R samples, represented in Green, and paths that were inconsistently clustered. A t-test detected a significant difference between the two distributions ($p < 0.0001$) indicating that paths that more tightly resembled the average trajectory of their corresponding bundle were more likely to be consistently clustered across populations, overall validating the consistency of our Behavioral-Tractography analysis across samples.
**Panel C:** Correlation of average 3D coordinates, characterizing the average trajectory of the 9 Behavioral-Tractography-Bundles across independent TD-D and TD-R sample.

**Supplementary Analysis 3: Replication analysis in a subsample of High-IQ 22q11DS individuals**

**Methods:** To verify that network differences observed across clinical samples were not entirely related to the higher prevalence of intellectual disability in the 22q11DS cohort, we conducted a supplementary analysis considering only individuals with Full-Scale-IQ > 70, described demographically in Supplementary Table 1. We compared the High-IQ-22q11DS sub-sample to ASD, with regards to the Connection Strength of individual connections, Connectivity-Dissimilarly-Index (CDI) of individual variables, average trajectory of Behavioral-Tractography Bundles and Behavioral Diffusion Analysis (BDA) of individual variables. We employed the same permutation-testing procedure described in the main text to test statistical significance of differences across samples which we describe in detail in Supplementary Table 1. Moreover, we correlated differences in the above-mentioned measures observed between High-IQ-22q11DS and ASD with those observed when comparing the entire samples of 22q11DS to ASD.

**Results:** The High-IQ-22q11DS sample was significantly different from the ASD sample in several of the above-mentioned metrics, including overall network structure captured by the average difference in connection strength of individual connections ($p<0.001$), CDI of multiple variables the trajectory of Behavioral-Tractography bundle 9 and BDA values of multiple variables in both Bundles 8 and 9. See Supplementary Table 2. Moreover the differences in the above-mentioned metrics measured across High-IQ-22q11DS and ASD samples strongly correlated with the differences observed across the entire samples described in the main text, with regards to differences in strength of individual connections ($R=0.7$, $p<0.0001$), CDI values (0.75, $p<0.0001$), Trajectories of Behavioral-Tractography-Bundles ($R=0.9$, $p<0.0001$) and BDA values of individual variables ($R=0.5$, $p<0.0001$). See Supplementary Figure 3.

**Discussion:** Overall, results show that network differences observed across 22q11DS and ASD were not significantly driven by 22q11DS with intellectual disability. Moreover, the High-IQ-22q11DS were on average younger than the entire 22q11DS sample, and not significantly different to the ASD sample. As such these supplementary analyses show that differences in age across 22q11DS and ASD samples were not significantly contributing to network differences described in the main text.

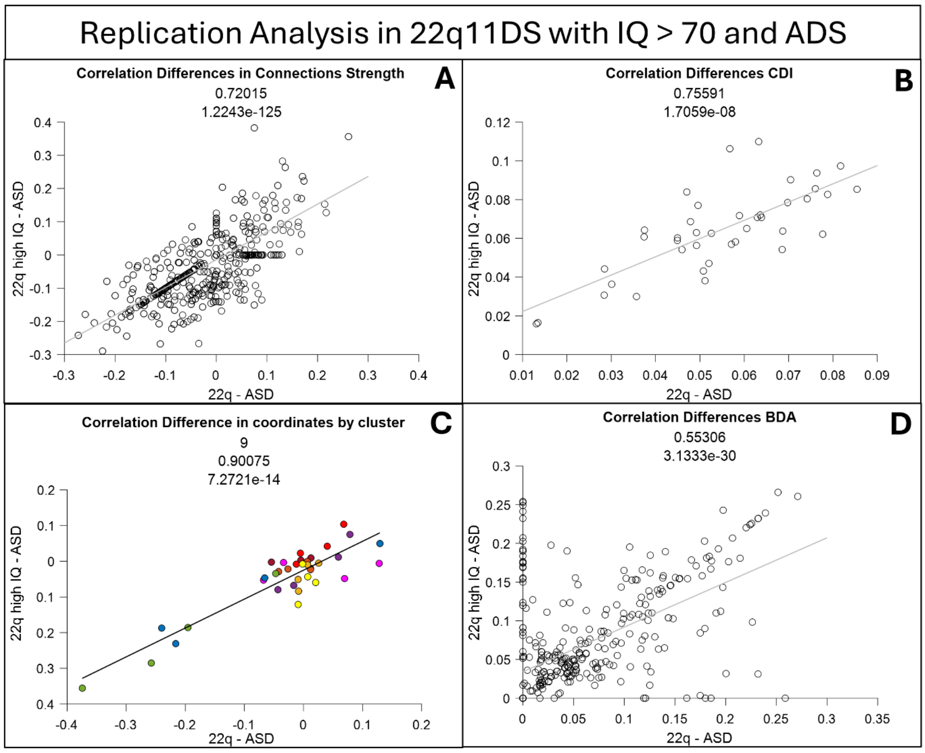


**Supplementary Figure 3:** Correlation of network differences measured in when comparing the entire sample of 22q11DS to ASD, plotted in the x axis and the subsample of High-IQ-22q11DS to the same ASD sample. **Panel A**: Correlation of differences in strength of individual network connections. **Panel B:** Correlation of differences in CDI **Panel C**: Correlation of differences in the average trajectory of Behavioral-Tractography bundles **Panel D:** Correlation of differences in B

**Supplementary Analysis 4: Stability and Replication in Group Differences in Behavioral Tractography Analysis**

To assess the robustness of BT cluster differences between populations, we performed a split-sample stability and replication analysis. Specifically, for each of 500 iterations, participants from the 22q11DS and ASD groups were randomly split into two equal halves. The first pair of 22q11DS and ASD subgroups served as the discovery samples while the second pair as the replication samples.

We estimated the consistency of differences in the trajectories of BT Bundles defined in the entire sample and described in the main text. We specifically computed differences in the 4 coordinates defining the trajectory of each BT Bundle (Exit TL1-x, Exit TL1-y, Entry TL2-x, Entry TL2-y), across 22q11DS and ASD Discovery and Replication Samples for each of 500 iterations.

We firstly estimated the consistency of group-differences across ASD and 22q11DS Discovery Samples, by calculating the proportion of iterations in which coordinated differences aligned with the direction observed in the entire sample. We compared this stability value against a null distribution derived by randomly assigning ASD and 22q11DS diagnostic labels across the 500 discovery samples for 1000 iterations. For each Bundle we also computed an overall consistency metric by multiplying differences observed in each of 4 coordinates with the direction of difference observed in the entire sample, and then adding such values across the 4 coordinates. We computed the proportion of Discovery samples in which such metric was positive, estimating overall consistency of trajectory differences for a given Bundle.

Next to estimate the replicability of group differences, we computed the proportion of iterations where concordant direction of group differences were observed across independent discovery and replication samples. We computed replicability metrics for each Bundle coordinate, as well as a global replicability metric derived by multiplying differences observed in each of 4 coordinates in 22q11DS vs ASD replication samples with the direction of difference observed in the Discovery Samples, and then adding such values across the 4 Bundle coordinates. We compared these values against a null distribution obtained by randomly assigning 22q11DS and ASD diagnostic labels across the 500 replication samples for 1000 iterations.

As a final analysis we repeated the entire procedure while re-running the clustering procedure employed to define BT Bundles for each iteration of discovery samples (Figure 4B).

**Results:**

Bundle 7 and 9 that were characterized by statistically significant differences in trajectory across the entire 22q11DS and ASD samples (see main text), also showed highest consistency of trajectory differences across ASD and 22q11DS discovery samples (Bundle-9 Consistency= 98%, $p<0.001$, Budle-7 Consistency= 92%, $p<0.0001$) Moreover Bundle 7 and 9 where the only Bundles to display high replicability of trajectory differences across Discovery and Replication samples (Bundle-9 Replicability = 98%, $p<0.0001$, Bundle-7 Replicability = 68%, $p=0.02$). See Supplementary Figure 4A for details.

As would be expected both consistency and replicability values were lower when modifying composition of BD Bundles at each Discovery Sample iteration. For Bundle-9 both consistency and replicability values remained statistically significant (Bundle-9 Consistency = 80%, $p<0.0001$, Bundle-9 Replicability = 74%, $p=0.02$), while for Bundle-7 only consistency values remain significant (Bundle-7 Consistency = 72%, $p<0.0001$, Bundle-7 Replicability = 46%, $p=0.8$). When modifying Bundle composition instead slightly increased Replicability of Bundle-4 trajectory differences (Bundle-7 Consistency = 88%, $p<0.0001$, Bundle-7 Replicability = 68%, $p=0.02$). See Supplementary Figure 4B for details.

**Discussion:**

The results of the analysis demonstrate that only the bundles exhibiting significant 3D trajectory bundle between-group differences (bundles7 and 9), also showed stability in the direction of these differences across discovery and replication samples. This finding suggests that the observed differences between the 22q and ASD groups are not contingent upon any particular data subset, but rather represent a consistent pattern that holds across various random partitions of the data.

In the second part of our analysis, where the clustering solution was recalculated independently for each discovery sample, we observed that, even when accounting for clustering variability, bundle 9 remained stable. This further reinforces the stability of the between-group differences, showing that the observed differences between the 22q and ASD groups are not reliant on a specific clustering configuration. Replicability in trajectory differences of Bundle-7 were instead more sensitive to variation in Bundle composition, implying that group differences described in Bundle-7 may be linked to a specific sub-set of pathways, that may be less consistently dissected as a single Bundle across discovery sample iterations.

**Supplementary Figure 4A: Stability in Group Differences in Behavioral Tractography Analysis**

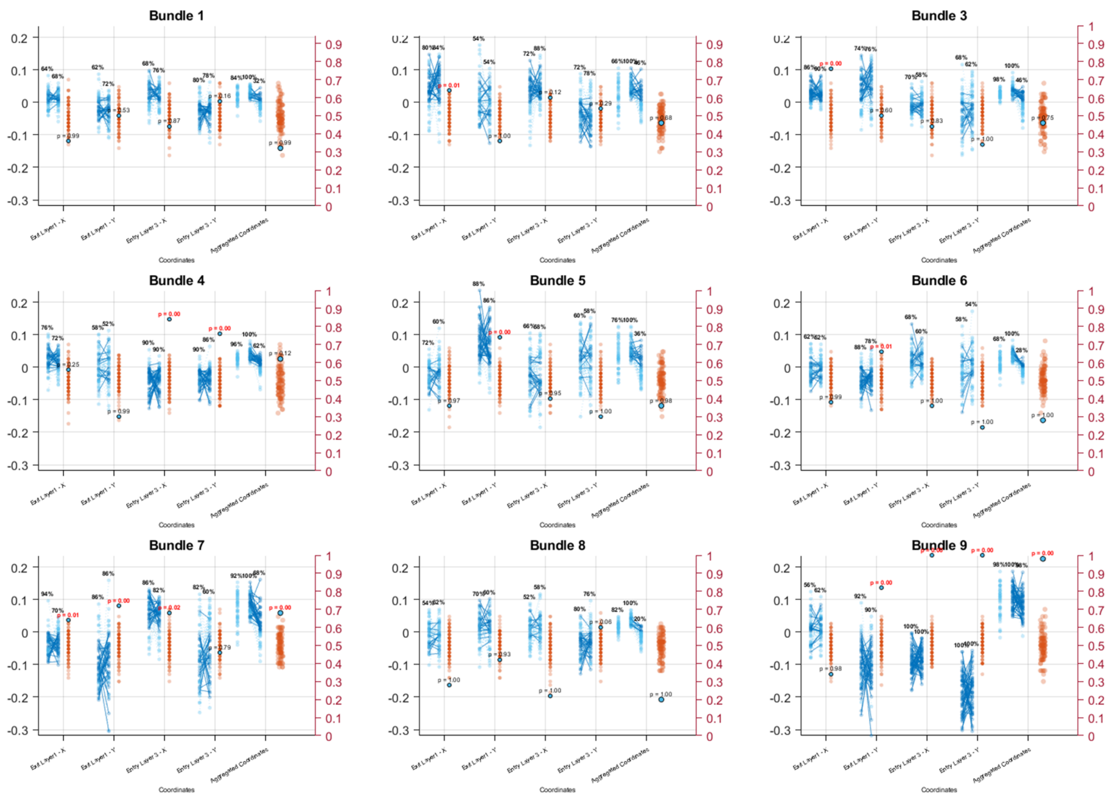


Each subplot corresponds to one bundle (n = 9). Within each subplot, five columns represent, from left to right: Exit TL1 - X, Exit TL1 - Y, Entry TL2 - X, Entry TL2 - Y, and the aggregate cluster-level measure. Each column is further subdivided into three sub-columns:

1. **Discovery sample** – distribution of 22q vs. ASD difference directions across 500 half-sample iterations.
2. **Replication sample** – corresponding differences in the replication sample; each dot represents one iteration, connected by lines to its discovery counterpart.
3. **Null distribution** – generated by randomly inverting the direction of replication differences; observed replication consistency is shown in blue, null values in red.

Percentage values above the first and second sub-columns indicate the proportion of iterations consistent with the direction of group differences observed in the entire sample, in the discovery and replication datasets respectively. P-values above the third sub-column indicate whether replication consistency significantly exceeded chance expectations.

In the fifth column (*aggregate cluster-level measure*), an additional sub-column is shown on the left ("consistency"), depicting the distribution and percentage of discovery iterations in which the *discovery consistency metric* was positive.

**Supplementary Figure 4B: Stability Group Differences in Behavioral Tractography Analysis**

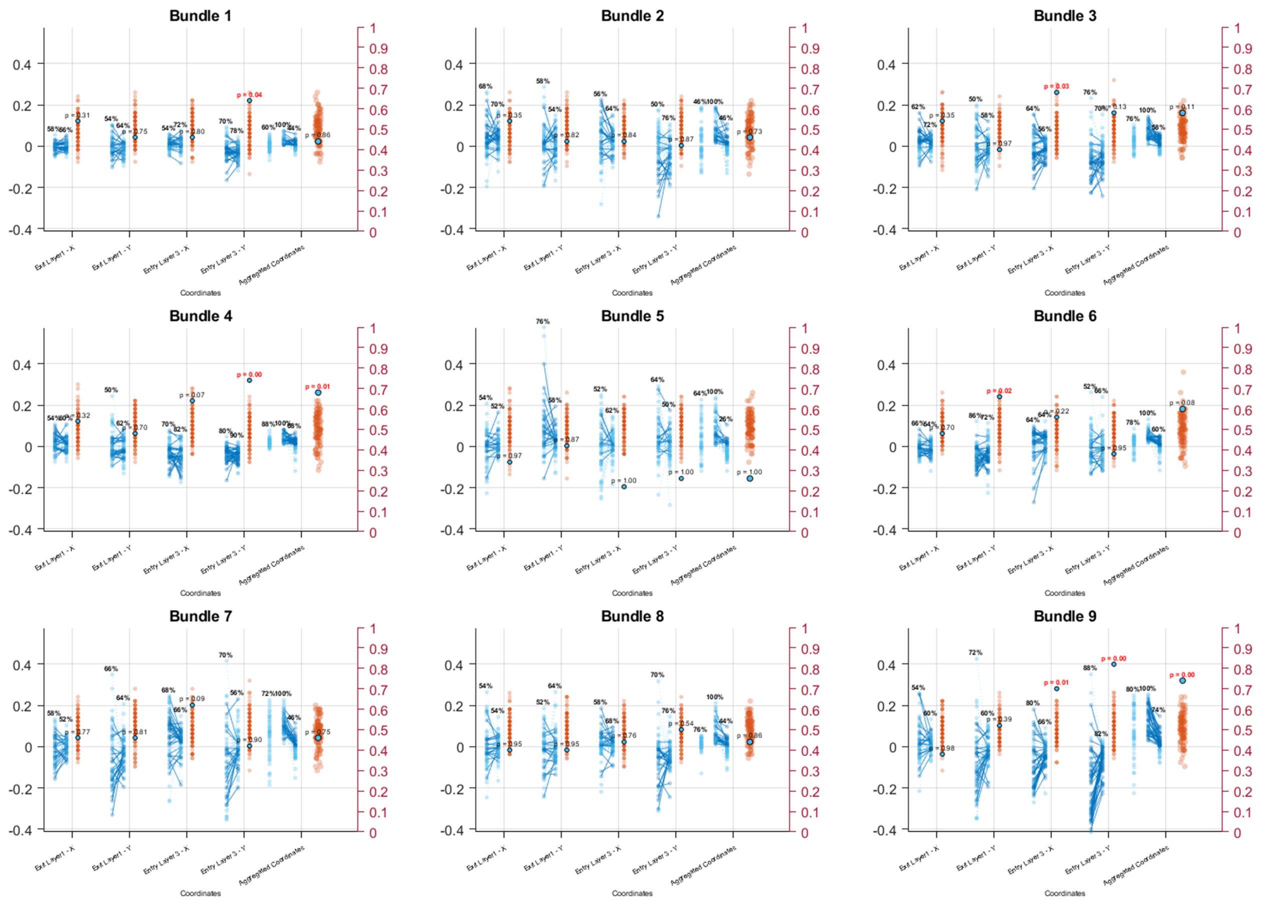


Same structure and layout as Figure 4, but cluster assignments were derived independently within each discovery sample at each iteration via K-means clustering. This approach allows assessing the robustness of between-group differences while accounting for potential variability in clustering solutions across subsamples.

| | TD-D (N=53) | 22q11DS High IQ (N=40) | ASD (N=60) | P-Value TD-D-ASD | P-Value HC-22q11DS High IQ | P-Value ASD-22q11DS High IQ |
|---|---|---|---|---|---|---|
| **Gender (M/F)** | 27/26 | 24/16 | 29/31 | p=0.78 | p=0.39 | p=0.26 |
| **Age** | 18.5±3.7 | 19±4.69 | 17.7±4.9 | p=0.37 | p=0.55 | p=0.21 |
| **Mean # of Assessments** | 29.4±8.8 | 27.45±11.02 | 30±9.4 | p=0.73 | p=0.34 | p=0.21 |
| **# of Consecutive Assessments** | 21.90±7.82 | 20.25±9.75 | 22.22±8.64 | p=0.84 | p=0.37 | p=0.29 |
| **Full Scale IQ** | 111.2±11.9 | 82.02±8.21 | 107.7±15.9 | p=0.20 | p<0.001 | P<0.001 |

**Supplementary Table 1:** Demographics data for Typically Development Discovery Sample (as in the main text), 22Q High IQ (>70), ASD (as in the main text). Gender differences were tested with Chi-Square test. Continuous variables were tested with Two-Sample T-Tests.

| CDI (22Q - ASD) | CONNECTIVITY DISSIMILARITY INDEX | | | | | |
|---|---|---|---|---|---|---|
| | 22q11DS vs ASD | | 22q11DS vsTD-D | | TD-D vs ASD | |
| SYMPTOMS | T1 | T2 | T1 | T2 | T1 | T2 |
| Lack_Relaxation | 0.02 | 0.00 | 0.11 | 0.22 | 0.06 | 0.01 |
| Loneliness | 0.27 | 0.32 | 0.31 | 0.34 | 0.10 | 0.08 |
| Anxiety | 0.03 | 0.01 | 0.97 | 0.88 | 0.03 | 0.01 |
| Lack_Happiness | 0.01 | 0.09 | 0.02 | 0.07 | 0.28 | 0.73 |
| Irritation | 0.27 | 0.29 | 0.13 | 0.17 | 0.05 | 0.11 |
| Sensory_Issue | 0.02 | 0.04 | 0.91 | 0.80 | 0.03 | 0.03 |
| Lack_Excitement | 0.08 | 0.06 | 0.13 | 0.06 | 0.84 | 0.52 |
| Sadness | 0.36 | 0.32 | 0.88 | 0.65 | 0.10 | 0.06 |
| Lack_Confidence | 0.32 | 0.15 | 0.89 | 0.53 | 0.95 | 0.88 |
| Feeling_Rejected | 0.48 | 0.61 | 0.80 | 0.66 | 0.27 | 0.04 |
| Feeling_Unsafe | 0.04 | 0.06 | 0.91 | 0.98 | 0.01 | 0.11 |
| Confusion | 0.29 | 0.22 | 0.58 | 0.80 | 0.51 | 0.45 |
| Hallucinations | 0.08 | 0.09 | 0.89 | 0.97 | 0.39 | 0.18 |
| Feeling_Tired | 0.03 | 0.01 | 0.00 | 0.00 | 0.32 | 0.31 |
| Lack_Motivation | 0.02 | 0.02 | 0.02 | 0.02 | 0.23 | 0.85 |
| Lack_Physical_Activity | 0.05 | 0.09 | 0.79 | 0.75 | 0.13 | 0.04 |
| Finding_Activity_Difficult | 0.02 | 0.03 | 0.66 | 0.18 | 0.10 | 0.15 |
| Lack_Enjoing_Actvity | 0.00 | 0.02 | 0.01 | 0.13 | 0.22 | 0.58 |
| Lack_Concentration | 0.65 | 0.19 | 0.11 | 0.05 | 0.08 | 0.17 |
| Being_Alone | 0.69 | 0.35 | 0.01 | 0.02 | 0.04 | 0.02 |

| BDA 22q - ASD | BEHAVIORAL DIFFUSION ANALYSIS | | | | | | | | | | | | | | | | | |
|---|---|---|---|---|---|---|---|---|---|---|---|---|---|---|---|---|---|---|
| Cluster- Bundle | 1 | | 2 | | 3 | | 4 | | 5 | | 6 | | 7 | | 8 | | 9 | |
| Number of Paths | 59 | | 41 | | 44 | | 48 | | 30 | | 62 | | 27 | | 60 | | 29 | |
| P-value BT | 0.97 | | 0.29 | | 0.25 | | 0.20 | | 0.16 | | 0.44 | | 0.00 | | 0.98 | | 0.00 | |
| SYMPTOMS | T1 | T2 | T1 | T2 | T1 | T2 | T1 | T2 | T1 | T2 | T1 | T2 | T1 | T2 | T1 | T2 | T1 | T2 |
| Lack_Relaxation | 0.67 | 0.79 | 1.00 | 0.32 | 0.04 | 0.12 | 0.17 | 1.00 | 0.92 | 0.36 | 1.00 | 0.05 | 0.11 | 1.00 | 0.18 | 0.65 | 1.00 | 0.02 |
| Loneliness | 1.00 | 0.87 | 0.07 | 0.96 | 1.00 | 1.00 | 0.97 | 0.74 | 0.02 | 1.00 | 1.00 | 0.97 | 0.07 | 0.04 | 0.95 | 0.80 | 0.04 | 0.05 |
| Anxiety | 0.70 | 0.84 | 1.00 | 0.52 | 0.58 | 0.58 | 0.66 | 1.00 | 0.97 | 0.04 | 1.00 | 0.62 | 1.00 | 0.41 | 0.97 | 0.24 | 0.09 | 0.16 |
| Lack_Happiness | 0.76 | 0.07 | 1.00 | 0.51 | 0.71 | 0.45 | 0.55 | 0.33 | 0.36 | 1.00 | 0.80 | 0.25 | 0.41 | 0.00 | 0.56 | 0.15 | 0.01 | 0.00 |
| Irritation | 0.60 | 0.57 | 0.03 | 0.69 | 0.95 | 1.00 | 0.53 | 1.00 | 0.21 | 0.07 | 1.00 | 0.46 | 1.00 | 0.42 | 0.02 | 0.65 | 0.21 | 0.50 |
| Sensory_Issue | 1.00 | 0.50 | 0.64 | 0.09 | 1.00 | 1.00 | 0.85 | 1.00 | 0.34 | 0.03 | 1.00 | 0.97 | 0.35 | 0.00 | 0.09 | 1.00 | 0.09 | 0.54 |
| Lack_Excitement | 1.00 | 1.00 | 1.00 | 0.41 | 0.72 | 0.72 | 0.42 | 0.21 | 1.00 | 1.00 | 0.80 | 0.45 | 0.11 | 1.00 | 1.00 | 0.28 | 1.00 | 0.00 |
| Sadness | 0.56 | 0.82 | 1.00 | 0.11 | 1.00 | 1.00 | 0.63 | 1.00 | 0.04 | 0.82 | 1.00 | 0.99 | 0.22 | 0.26 | 0.84 | 0.98 | 0.00 | 0.01 |
| Lack_Confidence | 0.85 | 0.66 | 1.00 | 0.51 | 0.34 | 0.77 | 0.72 | 0.17 | 0.47 | 1.00 | 1.00 | 0.75 | 1.00 | 0.70 | 1.00 | 0.08 | 0.58 | 0.51 |
| Feeling_Rejected | 0.25 | 0.05 | 0.48 | 0.68 | 0.02 | 1.00 | 0.14 | 1.00 | 0.09 | 0.79 | 1.00 | 0.88 | 1.00 | 0.66 | 0.79 | 0.86 | 0.00 | 1.00 |
| Feeling_Unsafe | 1.00 | 1.00 | 0.22 | 0.12 | 0.48 | 1.00 | 0.51 | 1.00 | 0.76 | 0.35 | 1.00 | 0.07 | 1.00 | 0.20 | 0.94 | 0.13 | 0.62 | 0.70 |
| Confusion | 1.00 | 0.27 | 0.89 | 0.43 | 0.89 | 1.00 | 0.89 | 0.86 | 1.00 | 0.43 | 1.00 | 1.00 | 1.00 | 0.19 | 0.51 | 1.00 | 1.00 | 1.00 |
| Hallucinations | 1.00 | 0.88 | 0.59 | 0.13 | 1.00 | 1.00 | 0.81 | 1.00 | 0.56 | 0.20 | 1.00 | 0.18 | 1.00 | 0.32 | 1.00 | 1.00 | 0.05 | 1.00 |
| Feeling_Tired | 1.00 | 1.00 | 1.00 | 0.05 | 0.11 | 0.11 | 0.79 | 0.63 | 1.00 | 1.00 | 0.09 | 0.05 | 0.59 | 0.23 | 0.78 | 0.28 | 0.39 | 0.22 |
| Lack_Motivation | 1.00 | 1.00 | 1.00 | 0.80 | 0.49 | 0.53 | 0.95 | 0.82 | 1.00 | 1.00 | 0.02 | 0.81 | 0.26 | 1.00 | 1.00 | 1.00 | 0.20 | 0.01 |
| Lack_Physical_Activity | 1.00 | 1.00 | 1.00 | 1.00 | 1.00 | 1.00 | 0.03 | 0.23 | 1.00 | 1.00 | 0.23 | 0.47 | 1.00 | 1.00 | 1.00 | 1.00 | 0.04 | 0.00 |
| Finding_Activity_Difficult | 0.17 | 0.25 | 1.00 | 0.08 | 1.00 | 0.51 | 0.21 | 0.14 | 1.00 | 1.00 | 0.20 | 0.62 | 0.00 | 0.67 | 0.03 | 0.46 | 0.00 | 0.67 |
| Lack_Enjoing_Actvity | 1.00 | 0.92 | 1.00 | 0.28 | 0.14 | 0.00 | 0.02 | 0.12 | 0.27 | 1.00 | 0.66 | 0.23 | 0.00 | 0.66 | 0.66 | 0.78 | 0.45 | 0.03 |
| Lack_Concentration | 1.00 | 1.00 | 1.00 | 0.36 | 1.00 | 0.05 | 0.01 | 0.08 | 0.08 | 1.00 | 0.96 | 0.23 | 0.04 | 1.00 | 1.00 | 1.00 | 0.17 | 0.01 |
| Being_Alone | 1.00 | 0.94 | 0.74 | 0.59 | 1.00 | 1.00 | 1.00 | 0.86 | 0.08 | 1.00 | 1.00 | 0.35 | 1.00 | 0.16 | 0.98 | 0.84 | 0.06 | 0.04 |

| CDI (22Q High IQ - ASD) | CONNECTIVITY DISSIMILARITY INDEX | | | | | |
|---|---|---|---|---|---|---|
| | 22q11DS high IQ vs ASD | | 22q11DS high IQvsTD-D | | TD-D vs ASD | |
| SYMPTOMS | T1 | T2 | T1 | T2 | T1 | T2 |
| Lack_Relaxation | 0.01 | 0.00 | 0.98 | 1.00 | 0.06 | 0.01 |
| Loneliness | 0.07 | 0.03 | 0.35 | 0.11 | 0.10 | 0.08 |
| Anxiety | 0.04 | 0.02 | 0.72 | 0.44 | 0.03 | 0.01 |
| Lack_Happiness | 0.01 | 0.03 | 0.27 | 0.21 | 0.28 | 0.73 |
| Irritation | 0.08 | 0.10 | 0.88 | 0.95 | 0.05 | 0.11 |
| Sensory_Issue | 0.01 | 0.06 | 0.39 | 0.35 | 0.03 | 0.03 |
| Lack_Excitement | 0.21 | 0.11 | 0.40 | 0.41 | 0.84 | 0.52 |
| Sadness | 0.00 | 0.00 | 0.95 | 0.92 | 0.10 | 0.06 |
| Lack_Confidence | 0.07 | 0.16 | 0.70 | 0.86 | 0.95 | 0.88 |
| Feeling_Rejected | 0.04 | 0.02 | 0.99 | 0.93 | 0.27 | 0.04 |
| Feeling_Unsafe | 0.01 | 0.07 | 0.32 | 0.89 | 0.01 | 0.11 |
| Confusion | 0.01 | 0.01 | 0.11 | 0.00 | 0.51 | 0.45 |
| Hallucinations | 0.01 | 0.00 | 0.50 | 0.64 | 0.39 | 0.18 |
| Feeling_Tired | 0.35 | 0.21 | 0.02 | 0.02 | 0.32 | 0.31 |
| Lack_Motivation | 0.00 | 0.03 | 0.10 | 0.00 | 0.23 | 0.85 |
| Lack_Physical_Activity | 0.06 | 0.00 | 0.25 | 0.52 | 0.13 | 0.04 |
| Finding_Activity_Difficult | 0.14 | 0.26 | 0.72 | 0.15 | 0.10 | 0.15 |
| Lack_Enjoing_Actvity | 0.03 | 0.11 | 0.00 | 0.07 | 0.22 | 0.58 |
| Lack_Concentration | 0.09 | 0.16 | 1.00 | 1.00 | 0.08 | 0.17 |
| Being_Alone | 0.70 | 0.42 | 0.00 | 0.00 | 0.04 | 0.02 |

| BDA 22q High IQ - ASD | BEHAVIORAL DIFFUSION ANALYSIS | | | | | | | | | | | | | | | | | |
|---|---|---|---|---|---|---|---|---|---|---|---|---|---|---|---|---|---|---|
| Cluster- Bundle | 1 | | 2 | | 3 | | 4 | | 5 | | 6 | | 7 | | 8 | | 9 | |
| Number of Paths | 55 | | 38 | | 35 | | 57 | | 34 | | 54 | | 57 | | 33 | | 37 | |
| P-value BT | 0.02 | | 0.34 | | 0.13 | | 0.57 | | 0.82 | | 0.93 | | 0.05 | | 0.02 | | 0.00 | |
| SYMPTOMS | T1 | T2 | T1 | T2 | T1 | T2 | T1 | T2 | T1 | T2 | T1 | T2 | T1 | T2 | T1 | T2 | T1 | T2 |
| Lack_Relaxation | 0.15 | 0.22 | 1.00 | 0.09 | 0.10 | 0.34 | 0.10 | 0.61 | 0.25 | 0.46 | 0.24 | 0.57 | 0.64 | 0.49 | 0.39 | 0.39 | 0.44 | 0.01 |
| Loneliness | 0.29 | 0.38 | 0.61 | 0.95 | 1.00 | 1.00 | 0.83 | 0.62 | 0.28 | 0.33 | 1.00 | 0.59 | 1.00 | 0.00 | 0.16 | 0.25 | 0.00 | 0.22 |
| Anxiety | 0.66 | 0.61 | 0.12 | 0.29 | 0.48 | 0.39 | 0.63 | 1.00 | 0.29 | 0.83 | 1.00 | 0.67 | 0.10 | 0.71 | 0.66 | 0.51 | 0.03 | 0.24 |
| Lack_Happiness | 0.91 | 0.27 | 1.00 | 0.25 | 0.55 | 0.05 | 0.23 | 0.28 | 0.03 | 1.00 | 0.83 | 0.72 | 0.77 | 0.00 | 0.43 | 0.37 | 0.00 | 0.28 |
| Irritation | 0.74 | 0.02 | 0.05 | 0.79 | 0.51 | 0.88 | 0.78 | 1.00 | 0.36 | 0.23 | 1.00 | 0.74 | 1.00 | 0.25 | 0.88 | 0.32 | 0.13 | 0.55 |
| Sensory_Issue | 0.42 | 0.44 | 0.45 | 0.46 | 1.00 | 1.00 | 0.67 | 1.00 | 0.90 | 0.97 | 1.00 | 1.00 | 0.46 | 0.00 | 0.13 | 0.44 | 0.02 | 0.46 |
| Lack_Excitement | 1.00 | 1.00 | 1.00 | 0.32 | 0.76 | 0.48 | 0.24 | 0.19 | 1.00 | 1.00 | 0.87 | 0.65 | 0.13 | 0.04 | 1.00 | 0.46 | 1.00 | 0.06 |
| Sadness | 0.02 | 0.00 | 0.60 | 0.87 | 1.00 | 0.02 | 0.51 | 1.00 | 0.01 | 0.59 | 1.00 | 0.92 | 0.44 | 0.40 | 0.32 | 0.00 | 0.01 | 0.01 |
| Lack_Confidence | 0.10 | 0.02 | 0.34 | 0.54 | 0.15 | 0.84 | 0.18 | 0.35 | 0.84 | 0.58 | 1.00 | 0.57 | 1.00 | 0.35 | 0.51 | 0.00 | 0.00 | 0.41 |
| Feeling_Rejected | 0.37 | 0.02 | 0.73 | 0.35 | 1.00 | 0.89 | 1.00 | 1.00 | 0.17 | 0.28 | 1.00 | 0.58 | 1.00 | 0.63 | 0.70 | 0.38 | 0.06 | 1.00 |
| Feeling_Unsafe | 1.00 | 0.43 | 0.07 | 0.77 | 0.64 | 1.00 | 0.23 | 1.00 | 0.29 | 0.71 | 1.00 | 0.46 | 1.00 | 0.56 | 0.43 | 0.66 | 0.00 | 0.51 |
| Confusion | 1.00 | 0.54 | 0.63 | 1.00 | 0.80 | 1.00 | 0.88 | 1.00 | 1.00 | 0.49 | 1.00 | 0.95 | 1.00 | 0.15 | 0.51 | 1.00 | 0.07 | 0.07 |
| Hallucinations | 0.01 | 0.01 | 0.54 | 0.08 | 1.00 | 1.00 | 0.76 | 1.00 | 1.00 | 0.50 | 1.00 | 0.74 | 1.00 | 0.32 | 1.00 | 1.00 | 0.04 | 1.00 |
| Feeling_Tired | 1.00 | 1.00 | 0.13 | 0.85 | 0.37 | 0.49 | 0.82 | 0.80 | 1.00 | 1.00 | 0.59 | 0.52 | 0.55 | 0.37 | 0.08 | 0.63 | 0.03 | 0.00 |
| Lack_Motivation | 0.06 | 1.00 | 1.00 | 0.80 | 0.45 | 0.72 | 0.40 | 0.79 | 0.00 | 1.00 | 0.65 | 0.42 | 0.29 | 1.00 | 1.00 | 0.00 | 0.07 | 0.00 |
| Lack_Physical_Activity | 1.00 | 1.00 | 1.00 | 1.00 | 1.00 | 1.00 | 0.00 | 0.25 | 1.00 | 1.00 | 0.17 | 0.35 | 1.00 | 1.00 | 0.03 | 1.00 | 0.05 | 0.00 |
| Finding_Activity_Difficult | 0.53 | 0.43 | 1.00 | 0.02 | 1.00 | 0.54 | 0.65 | 0.70 | 0.01 | 1.00 | 0.35 | 0.90 | 0.00 | 0.77 | 0.00 | 0.14 | 0.00 | 0.79 |
| Lack_Enjoing_Actvity | 1.00 | 0.54 | 1.00 | 0.77 | 0.72 | 0.00 | 0.58 | 0.44 | 0.11 | 0.05 | 0.74 | 0.26 | 0.00 | 0.75 | 0.43 | 0.87 | 0.15 | 0.00 |
| Lack_Concentration | 1.00 | 1.00 | 1.00 | 0.02 | 1.00 | 0.13 | 0.17 | 0.44 | 0.83 | 1.00 | 0.92 | 1.00 | 0.11 | 1.00 | 1.00 | 1.00 | 1.00 | 0.00 |
| Being_Alone | 0.15 | 0.45 | 0.85 | 0.85 | 1.00 | 1.00 | 0.09 | 0.87 | 0.00 | 0.21 | 1.00 | 0.09 | 1.00 | 0.00 | 0.13 | 0.15 | 0.00 | 0.10 |

### Supplementary Table 2:

The table presents results from the Connectivity Dissimilarity Index (CDI) and Behavioral Diffusion Analysis (BDA). It is organized into two main sections. The left section displays the p-values for CDI comparisons across the three groups (22q11DS. TD-D, and ASD). The right section provides, for each bundle (columns 1 to 9), the number of shortest paths included in the bundle, the p-value reflecting clinical differences in behavioral pathways identified by Behavioral Tractography, and the p-values from the Behavioral Diffusion Analysis, separated into t1 (temporal layer 1) and t2 (temporal layer 2). The lower part of the table repeats this structure, but includes only 22q11DS participants with IQ greater than 70.

### Ema Protocol

#### General Informations

- 8 notifications per day between 7:30 a.m. and 10:00 p.m. for 6 days
- At least 30 minutes between two notifications
- 15-minute window to respond before the notification disappears
- 3 blocks of questions (mood/symptomatology, context, event)

#### EMA Items

All EMA items were originally written in French and are presented here translated into English. Items shown in bold indicate those selected for analysis in this paper.

- **Mood/Symptoms**
    - **Right now, I feel relaxed (var = Lack Relaxation)**
    - **Right now, I feel lonely (var = Loneliness)**
    - **Right now, I feel anxious (var = Anxiety)**
    - **Right now, I feel happy, joyful (var = Lack Happiness)**
    - **Right now, I feel irritated, angry (var = Irritation)**
    - **Right now, I feel bothered by sensory stimulation (var = Sensory Issue)**
    - **Right now, I feel excited (var = Lack Excitement)**
    - **Right now, I feel sad (var = Sadness)**
    - **Right now, I feel confident (var = Lack Confidence)**
    - **Right now, I feel like others don't like me (var = Feeling Rejected)**
    - **Right now, I feel like I have to stay on guard, that I'm not safe (var = Feeling Unsafe)**
    - **Right now, I feel like my imagination is blending with reality (var = Confusion)**
    - **Right now, I feel like I'm hearing or seeing things that others don't perceive (var = Hallucinations)**
    - **Right now, I feel tired (var = Feeling Tired)**
    - **Right now, I feel like doing many things, I feel motivated (var = Lack Motivation)**
    - **Since the last beep, I have been physically active (var = Lack Physical Activity)**

- **Contexte**
    - What are you doing? (just before the beep) (var = Cur_activ)
    - **This activity is difficult (var = Finding Activity Difficult)**
    - **I am enjoying this activity (var = Lack Enjoying Activity)**
    - **I am focused on this activity (var = Lack Concentration)**
    - Where are you? (juste avant le beep) (var = Location)
    - **Are you alone? (var = Being Alone)**
    - Yes (=1)
        - I would prefer to be with other people (var = Pref_other)
        - I like being alone (var = Likealone)
        - I feel excluded, rejected (var = Feel_exclu)
    - No (=0)
        - Who are you with?
        - I would prefer to be alone (var = Pref_alone)
        - This company is pleasant (var = likecomp)
        - We are doing something together (var = interact)
        - I feel judged by this/these person(s) (var = feel_judged)
        - I am nervous in the presence of this/these person(s) (var = feel_nerv)
- Evénement
    - Now think about the most important event that has happened since the last beep
    - This event was pleasant
    - This event was stressful

- This event was important
- Were you alone during this event?
- Now think about the most important event that will happen in the next hour
- I am looking forward to this event
- In which category does this event fall?
- Will you be alone during this event?

**Response scale:**
1 (not at all), 2 (very slightly), 3 (slightly), 4 (moderately), 5 (strongly), 6 (very strongly), 7 (extremely)
**Context: activity:**
10 (work/school), 20 (household tasks), 30 (eating or drinking), 40 (personal care), 50 (rest/relaxation), 60 (social contact), 65 (online social contact), 70 (sports), 75 (TV, internet), 79 (other leisure), 89 (something else), 0 (nothing)
**Context: location:**
10 (home), 20 (school/work), 30 (at a friend's place), 40 (at a family member's place), 50 (hospital or care facility), 60 (public place), 70 (in transit), 89 (elsewhere)
**Context: with whom:**
10 (person living with me), 20 (person not living with me), 30 (boyfriend/girlfriend, spouse), 40 (friend), 50 (classmate), 60 (healthcare professional), 70 (acquaintance), 80 (pet), 89 (stranger), 99 (no one else)
**Event:**
0 (nothing), 10 (physical activity), 20 (work/school), 30 (leisure), 40 (sleeping), 50 (eating/drinking), 60 (medical activity), 89 (other)
